# Enhancing Fairness in Diabetes Prediction Systems through Smart User Interface Design

**DOI:** 10.1101/2025.06.04.25328959

**Authors:** Asma Tavangar, Zahra Arefzadeh, Ahmad Navid Asghari

## Abstract

**Objectives:** Artificial intelligence (AI) in chronic disease prediction often exhibits algorithmic biases, hindering equitable healthcare delivery. This study aims to develop and evaluate a Smart User Interface (Smart UI) framework that enhances fairness in diabetes prediction systems by operationalizing fairness at the human-computer interaction level, a dimension frequently overlooked in AI fairness research.

**Materials and Methods:** We employed a nine-metric fairness evaluation framework across four demographically diverse diabetes datasets (Kaggle, Pima Indian, Azure Open, CDC Health Indicators). The Smart UI integrates contextual adjustment tools, dynamic visualizations, real-time alerts, and transparent reporting, combining structured EHR data, wearable sensor inputs, and unstructured clinical notes via natural language processing. The framework was evaluated on a clinical dataset to assess fairness and performance improvements.

**Results:** The Smart UI significantly reduced disparities: for age, the equal opportunity difference (EOD) improved from 0.35 to 0.25, with accuracy rising from 90.52% to 91.83%; for BMI, EOD decreased from 0.56 to 0.38, with the F1-score increasing from 83.89% to 86.37%. These outcomes highlight the framework’s ability to enhance fairness without altering underlying algorithms.

**Discussion:** While the Smart UI demonstrates promise as a model-agnostic, scalable solution for equitable AI deployment, challenges such as data privacy, usability, and real-time processing persist. The framework’s reliance on diverse data sources and user-centered design underscores its potential, though validation in broader clinical settings is needed.

**Conclusion:** The Smart UI offers a replicable blueprint for embedding fairness in healthcare AI through interface design. Future research should focus on multicenter trials and applications to other chronic diseases to advance inclusive digital health solutions.

## 1 Introduction

Diabetes mellitus is one of the most prevalent chronic metabolic disorders worldwide, currently affecting over 537 million adults, with projections estimating a rise to more than 783 million by 2045 [1]. This growing burden is exacerbated by structural inequities in healthcare systems, which disproportionately impact underserved populations such as low-income communities [2], Indigenous groups [3], and women [4]. In recent years, artificial intelligence (AI) and machine learning (ML) have emerged as powerful tools for early disease prediction, enabling complex data analysis and personalized interventions in clinical settings [5].

Despite these advancements, integrating AI into real-world healthcare remains challenging, with algorithmic bias being a primary concern [6]. Many predictive models rely heavily on socioeconomic proxies—such as income, education level, geographic region, gender, or age—instead of direct biological indicators. This reliance can significantly increase error rates among vulnerable subpopulations, leading to unequal diagnostic outcomes for women, the elderly, and rural or Indigenous communities [6]. While previous efforts to mitigate bias have focused primarily on algorithmic refinements or data rebalancing [7], less attention has been given to the user interface—where data are entered, interpreted, and acted upon—and its critical role in detecting and addressing bias.

To address this gap, we propose a novel Smart User Interface (Smart UI) framework designed with three distinct user views: clinicians, patients, and researchers. Unlike existing fairness visualization tools such as Fairlearn and Aequitas—which are primarily research-oriented and emphasize post hoc bias analysis—our Smart UI is purposefully built to integrate with electronic health records (EHRs) and support real-time clinical decision-making. By providing dynamic alerts, contextual feature explanations, and patient-centric summaries, our system enhances utility across a broad spectrum of stakeholders and elevates fairness interventions beyond static analytics.

This framework uniquely integrates multi-metric fairness analysis into interactive interface design. It offers targeted interventions across four levels: contextual adjustment tools for high-risk features, dynamic risk visualization for medium-risk features, tailored alerts for low-risk predictors, and a results reporting system to enhance transparency and stakeholder engagement. The system allows users to explore the relative importance of features in model predictions and assess model performance across sensitive subgroups, such as women and older adults. This transparency fosters a deeper understanding of model behavior and enables real-time, human-in-the-loop adjustments to mitigate potential inequities. The framework also addresses implementation challenges, such as data privacy, usability, and real-time processing, through solutions like robust encryption, user-centered design, and scalable cloud platforms.

The framework was developed and evaluated using four publicly available, demographically diverse datasets: the Azure Open Diabetes Dataset, CDC Health Indicators, the Pima Indian Diabetes Dataset, and the Kaggle Diabetes Prediction Dataset. The process began by identifying potentially sensitive features through a combination of domain expertise and random forest feature importance analysis. A comprehensive nine-metric fairness evaluation framework, including performance disparities (e.g., accuracy differences) and group fairness metrics (e.g., equal opportunity difference), was applied to systematically assess and rank each feature’s equity risk. To reduce reliance on static and manual data entry, the interface integrates natural language processing (NLP) to extract insights from clinical notes.

Findings reveal significant disparities in model performance across several biological and demographic features. Attributes such as age, body mass index (BMI), glucose measures, insulin, gender, general health indicators, and familial diabetes history—though clinically relevant—were associated with notable disparities across datasets and subgroups. This highlights a critical insight: even scientifically valid clinical indicators can become sources of unintended bias when deployed without context-awareness (e.g., social or environmental factors). The proposed Smart UI framework not only identifies these inequities but also empowers users to understand and manage them through layered, interpretable interventions. By combining rigorous statistical analysis with transparent, user-centric design, the framework offers a practical, ethical, and scalable solution for achieving fairness in AI-driven diabetes care.

### Methods

This section delineates the methodological framework underpinning our fairness-aware diabetes prediction system, encompassing study design, data preprocessing, fairness evaluation metrics, and the development of the Smart User Interface (Smart UI). By integrating algorithmic fairness principles with user-centered design, our approach aims to mitigate bias and promote equitable outcomes across diverse patient populations—particularly in terms of prediction accuracy, treatment opportunity, and risk stratification.

A summary of the workflow is depicted in Figure 1, illustrating the key components and their interrelations, which are discussed in detail throughout the following subsections.

**Fig. 1.**
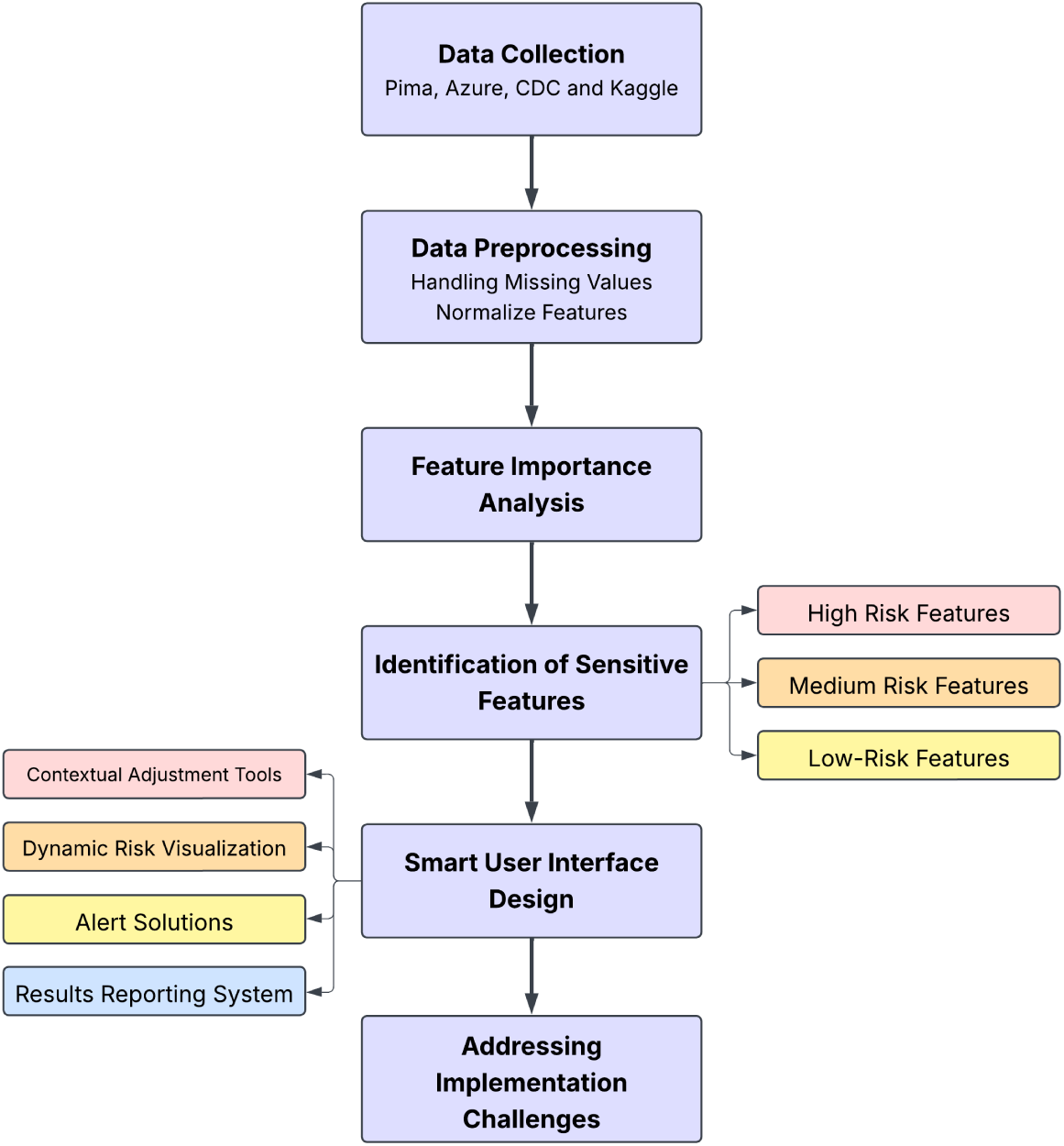
Overview of the methodological framework for fairness-aware diabetes prediction, highlighting key components and their interactions.

#### Study Design Overview

This study employed a retrospective, cross-sectional design integrating algorithmic fairness evaluation with user interface innovation. We analyzed four publicly available diabetes datasets—the Kaggle Diabetes Prediction Dataset, Pima Indian Diabetes Dataset, Azure Open Diabetes Dataset, and CDC Diabetes Health Indicators Dataset—to identify clinical, demographic, and socioeconomic features associated with predictive disparities. Each dataset was processed and modeled independently using a standardized machine learning pipeline, enabling consistent fairness assessments across diverse population samples.

Following the identification of bias-prone features using feature importance scores and a nine-criterion fairness evaluation framework, we stratified features into High, Medium, and Low Risk categories based on the magnitude and consistency of observed disparities. Building on these findings, we developed a Smart User Interface (UI) framework designed to mitigate algorithmic bias through contextual data adjustment, dynamic risk visualization, and fairness-aware alerts. The UI incorporates data from electronic health records, wearable devices, and natural language inputs to support transparency, usability, and equitable decision-making in diabetes prediction systems. It is worth noting that although the Pima Indian Diabetes Dataset includes only female participants, it provided a valuable opportunity to analyze the impact of pregnancy as a predictor—a feature often absent in other datasets but crucial for fairness analysis in gender-sensitive health outcomes.

#### Dataset Overview

This study draws upon four publicly available datasets to support the analysis of fairness in diabetes prediction systems: the Kaggle Diabetes Prediction Dataset [8], the Pima Indian Diabetes Dataset [9], the Azure Open Diabetes Dataset [10], and the CDC Diabetes Health Indicators Dataset [11]. These datasets were selected for their complementary strengths, including demographic heterogeneity, clinical richness, and socioeconomic variability. Such diversity is essential for detecting hidden biases and promoting generalizability in AI models deployed across real-world clinical populations.

Each dataset contributes unique perspectives: the Kaggle dataset emphasizes behavioral and lifestyle factors; the Azure dataset includes laboratory and biometric indicators; the CDC dataset captures a wide range of public health and socioeconomic variables; and the Pima dataset, though limited to female participants, provides a valuable lens into gender-specific factors such as pregnancy.

The varied feature spaces across datasets enable a multi-dimensional assessment of fairness, encompassing clinical, behavioral, and social determinants. Table 1 summarizes the key characteristics of each dataset, including sample size, target population, notable attributes, and relevance to fairness evaluation, offering a comparative reference for interpreting subsequent findings.

**Table 1.** Summary of Datasets Used in Fairness Analysis.

| Dataset | Size | Key Features | Total Number of Features | Fairness Relevance |
| --- | --- | --- | --- | --- |
| Kaggle Diabetes Prediction Dataset [8] | 10000 instances | Sex, Age, Hypertension, Heart Disease, Smoking History, BMI, HbA1c Level, Blood Glucose Level | 8 | Includes sex, age, and socioeconomic proxies (e.g., hypertension prevalence), enabling bias analysis across these axes. |
| Pima Indian Diabetes Dataset [9] | 768 instances | Number of Pregnancies, Plasma Glucose Concentration, Diastolic Blood Pressure, Triceps Skin Fold Thickness, 2-hour Serum Insulin Level, BMI, Diabetes Pedigree Function, Age | 8 | Homogeneous population (all female, single ethnic group) restricts generalizability; fairness limited to age and pregnancy-related bias analysis. |
| Azure Open Diabetes Dataset [10] | 442 instances | Age, Sex, BMI, Diastolic Blood Pressure, Total Cholesterol Ratio, LDL, HDL, Triglycerides, Long-term Glucose, Glucose Level in the Blood | 10 | Small size and unspecified demographics hinder fairness evaluation; time-series focus offers potential for individual-level bias tracking but lacks population diversity. |
| CDC Diabetes Health Indicators Dataset [11] | 25000 instances | BMI, High Blood Pressure, Age, Sex, Income, Physical Activity, General Health | 16 | Large, diverse sample supports fairness analysis across sex, age, income, and education; self-reported data may introduce bias, but broad coverage enhances equity insights. |

**Table 2.** Fairness criteria used to identify sensitive features.

| Criterion | Threshold | Fairness Relevance |
| --- | --- | --- |
| Regression Significance | $p < 0.01$ (Bonferroni-corrected) | Controls for multiple testing; reduces false positives |
| Accuracy Sensitivity | $\geq 3\%$ difference between groups | Indicates clinically meaningful disparities |
| AUC Sensitivity | $\geq 4\%$ difference between groups | Highlights reliability gaps in diagnostic predictions |
| F1-Score Sensitivity | $\geq 5\%$ difference between groups | Captures imbalance in classification effectiveness |
| Statistical Parity Difference (SPD) | $> 0.10$ or $< -0.10$ | Measures group-based disparity in positive outcomes |
| Disparate Impact (DI) | $< 0.80$ or $> 1.25$ | Reflects unequal treatment likelihood |
| Equal Opportunity Difference (EOD) | $> 0.10$ | Assesses true-positive-rate differences |
| Equalized Odds (EO) | $> 0.10$ | Evaluates fairness across both false positives and negatives |
| Odds Ratio (OR) | $< 0.75$ or $> 1.5$ | Epidemiologic benchmark for systemic bias |

Together, these datasets offer a robust foundation for our fairness analysis, capturing a wide range of clinical, demographic, and socioeconomic factors. Their combined diversity ensures that our study addresses biases across ethnic groups, socioeconomic strata, and clinical profiles, laying the groundwork for a generalizable and equitable diabetes prediction framework. This comprehensive dataset overview sets the stage for the subsequent evaluation of fairness metrics and the development of a Smart UI Design tailored to diverse clinical needs.

#### Preprocessing

To prepare the datasets for fairness analysis and predictive modeling, we applied preprocessing steps that balanced statistical rigor with equity-oriented considerations. For continuous variables (e.g., blood glucose, BMI), missing or anomalous values were imputed using the column mean. For categorical variables (e.g., smoking status, cholesterol check), missing values were filled using the mode (most frequent value), in order to prevent model overfitting to infrequent or underrepresented categories.

Following imputation, continuous features were normalized to the [0, 1] range using Min-Max Scaling, as shown in Equation 1, to prevent scale dominance during model training and to ensure comparability across heterogeneous datasets.

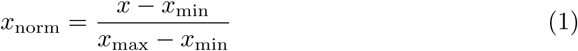

Subsequently, categorical variables—including income level, smoking history, and general health—were one-hot encoded to preserve subgroup granularity essential for downstream fairness analysis. To support consistent evaluation across datasets, feature names and category labels were harmonized to align semantically similar variables (e.g., GenHlth vs. General Health) and ensure interoperability.

These preprocessing steps were carefully designed to balance model readiness with fairness fidelity, ensuring that transformations did not obscure subgroup-specific disparities or dilute bias signals relevant to equity assessment.

#### Feature Importance Analysis

As a first step, we trained independent Random Forest classifiers on each of the four datasets to estimate the relative importance of features using Gini impurity scores. Random Forests were chosen due to their ability to capture non-linear relationships, model feature interactions, and produce robust importance estimates across heterogeneous data distributions.

Across all datasets, features such as Age, Body Mass Index (BMI), blood glucose indicators, and Diastolic Blood Pressure (DBP) consistently emerged as top predictors. To enhance clinical validity, these top-ranked features were cross-referenced with domain knowledge from endocrinology literature and public health guidelines [12–15].

This dual-filtering strategy—combining data-driven feature ranking with expert-informed validation—enabled us to construct a refined shortlist of potentially sensitive features.

Following this step, each feature in the shortlist was evaluated using a comprehensive nine-criterion fairness framework. This framework assessed feature influence from both a model performance and fairness perspective. A feature was classified as sensitive if it exceeded predefined thresholds in at least three of the nine criteria. This approach ensured that only features with multidimensional and statistically meaningful evidence of bias were retained for mitigation.

The fairness evaluation framework integrated multiple dimensions, including performance disparities (e.g., differences in accuracy, AUC, and F1-score), group fairness metrics (e.g., SPD, DI, EO), and statistical significance tests such as Regression Significance. This hybrid structure allowed us to balance model utility and equity considerations, ensuring that features contributing meaningfully to predictive accuracy were not inadvertently excluded, while also preventing bias amplification.

Rather than applying fairness metrics indiscriminately across all features, we focused our analysis on the most impactful and socially relevant predictors. Prior literature in fair machine learning [16–18] emphasizes that most algorithmic bias arises from a small subset of influential features. Comprehensive fairness analysis on weak predictors may lead to spurious findings, increased computational complexity, and dilution of truly actionable disparities.

Based on this hybrid framework, features were stratified into High, Medium, and Low Risk tiers, as visualized in Figure 2. This stratification served as the foundation for subsequent user interface design and bias mitigation strategies tailored to each risk level.

**Fig. 2.**
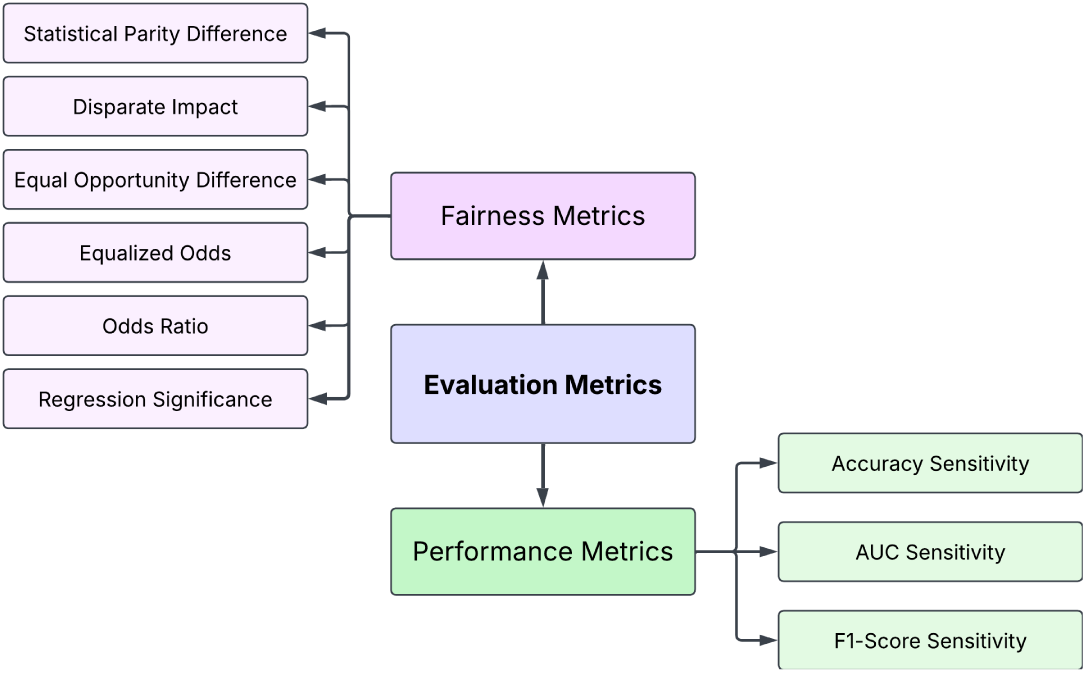
Categorization of sensitive features into risk tiers based on a hybrid framework combining fairness and performance metrics.

To estimate model performance and compute fairness metrics, we trained a Random Forest classifier with 100 estimators using an 80/20 train-test split, repeated across ten independent runs. Results were averaged, and standard deviations were reported to account for variability due to data partitioning. This design ensured reproducibility and statistical reliability in our fairness assessments.

#### Smart User Interface Design

Algorithmic bias in diabetes prediction systems can disproportionately affect vulnerable populations—such as older adults, women, and certain ethnic groups—resulting in unequal treatment outcomes. To address these disparities, we developed a Smart User Interface (Smart UI) framework that systematically embeds fairness principles into the user experience. This framework comprises four key components: Contextual Adjustment Tools, Dynamic Risk Visualization, Interactive Alert Solutions, and a Results Reporting System. Each component is designed to enhance transparency, interpretability, and equitable decision-making in clinical settings.

Unlike conventional interfaces that passively display model outputs, the Smart UI acts as an active decision-support agent. It dynamically adapts to clinical contexts, highlights fairness-related risks, and delivers tailored insights to users. These components align with trustworthy AI principles for fairness, usability, and clinical relevance [19, 20], proactively addressing algorithmic bias and promoting fair, transparent predictions. Through this design, the interface evolves from a passive output medium into an engaged, interactive partner in clinical decision-making.

#### Core Components

The Smart UI integrates a set of targeted components, each aligned with the level of risk associated with input features, to mitigate algorithmic bias in real-time. These components are designed to engage users actively in interpreting and evaluating model predictions, fostering more informed and fair clinical decisions. Figure 3 provides a visual overview of the UI architecture.

**Fig. 3.**
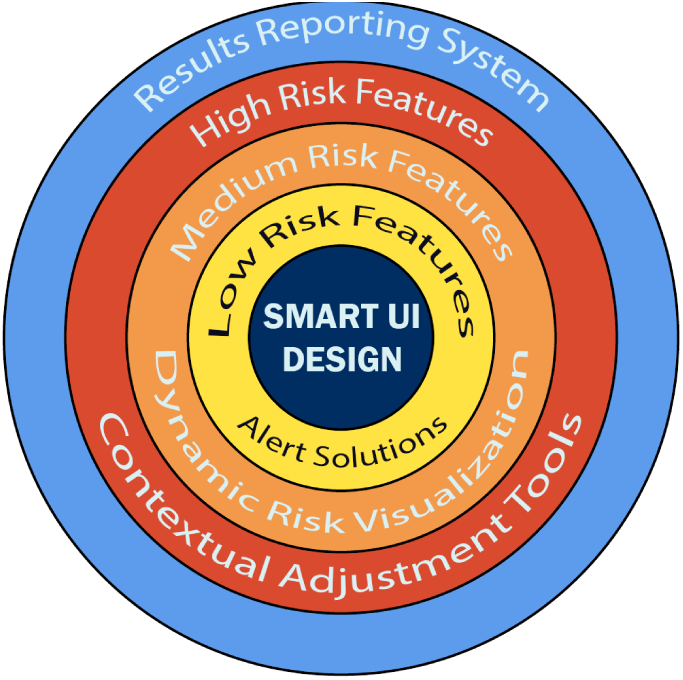
Core components of the Smart UI framework for fairness-aware diabetes prediction, organized by functional challenge areas.

**Fig. 4.**
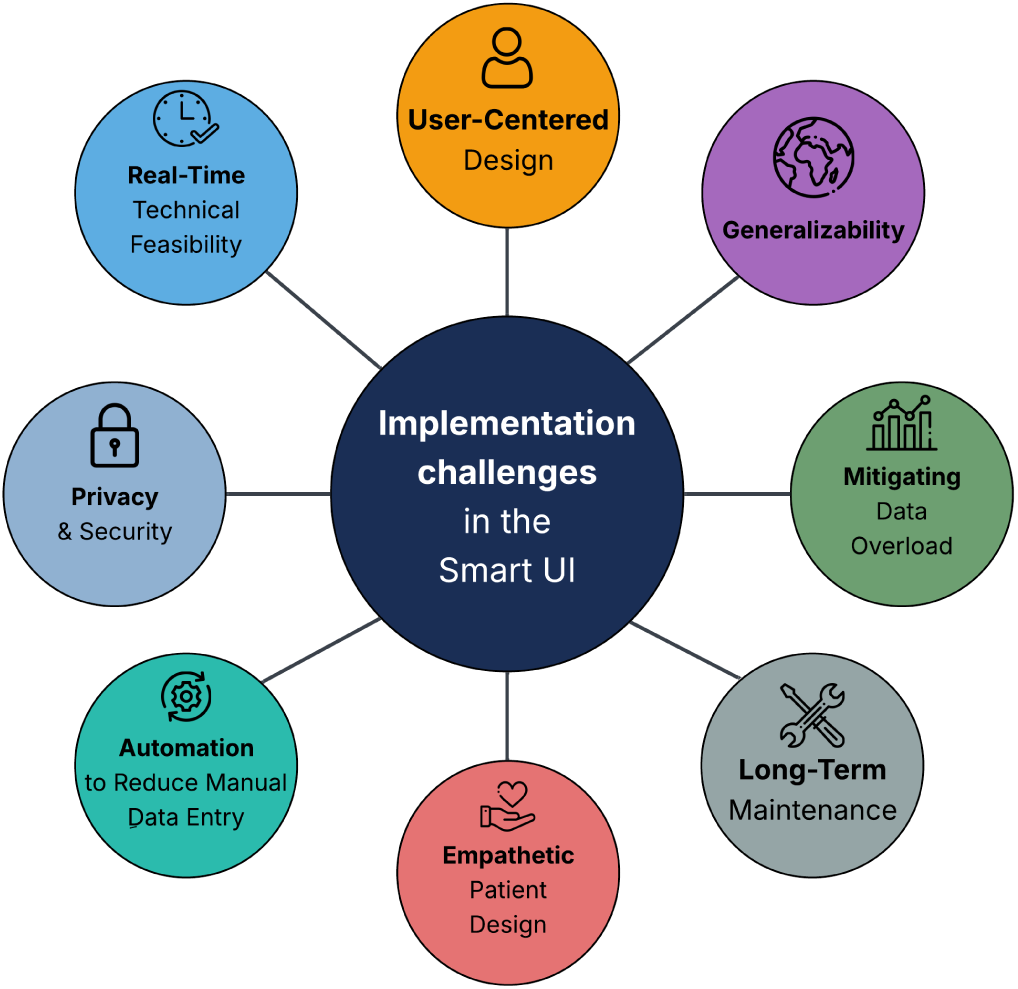
Key implementation challenges in the proposed Smart UI framework for fair diabetes prediction.

For features identified as high-risk—those with disproportionate influence on predictive outcomes—contextual adjustment tools are provided. These tools allow users to apply clinically and demographically sensitive calibrations, thereby moderating the feature’s impact and supporting a more equitable interpretation.

Medium-risk features are handled through dynamic risk visualization. This module uses interactive plots to illustrate how predicted risk evolves in response to different feature values, enabling deeper understanding of the model’s behavior across diverse patient subgroups.

Low-risk features, while generally stable, may still introduce unintended disparities under specific conditions. In such cases, the interface deploys real-time alert mechanisms—textual or color-coded—to draw attention to potential fairness violations or anomalous prediction patterns.

Finally, the results reporting system offers a structured and transparent presentation of model outputs, including both performance and fairness metrics. This module facilitates subgroup-based comparisons and equips clinicians and researchers with actionable information to evaluate model behavior through a fairness-aware lens.

##### High-Risk Features: Contextual Adjustment Tools

Features that violated all nine fairness criteria were classified as high-risk. In addition, features with fewer violations (e.g., six or seven) but exhibiting substantial disparities in performance—such as a 20% accuracy gap or an F1-score of 40% for certain subgroups—were also categorized as high-risk due to their potential for severe algorithmic bias. These features warrant focused intervention, as they may disproportionately affect predictive equity across vulnerable populations.

To mitigate such risks, the Smart User Interface (Smart UI) incorporates *Contextual Adjustment Tools*. These components enable users to input supplementary contextual data—such as dietary habits, physical activity, and sleep patterns—to refine model predictions in real time. The goal is to counterbalance the undue influence of high-risk features and improve predictive accuracy among underrepresented groups. This approach is supported by a substantial body of evidence showing that integrating behavioral and social determinants into machine learning models enhances both accuracy and fairness [13, 21, 22]. Specifically, studies in diabetes prediction have demonstrated that models enriched with contextual data yield better performance for marginalized populations and reduce systematic bias [23].

Table 3 presents a set of contextual adjustment strategies designed to mitigate bias associated with high-risk features in diabetes prediction. These strategies are grounded in behavioral, physiological, and socioeconomic data sources, aiming to improve predictive accuracy for underrepresented populations. The contextual inputs include information on physical activity, dietary intake, sleep patterns, psychological state, socioeconomic conditions, and medication usage—collected through various modalities such as wearable sensors, electronic health records (EHRs), food logs, and patient surveys.

**Table 3.**
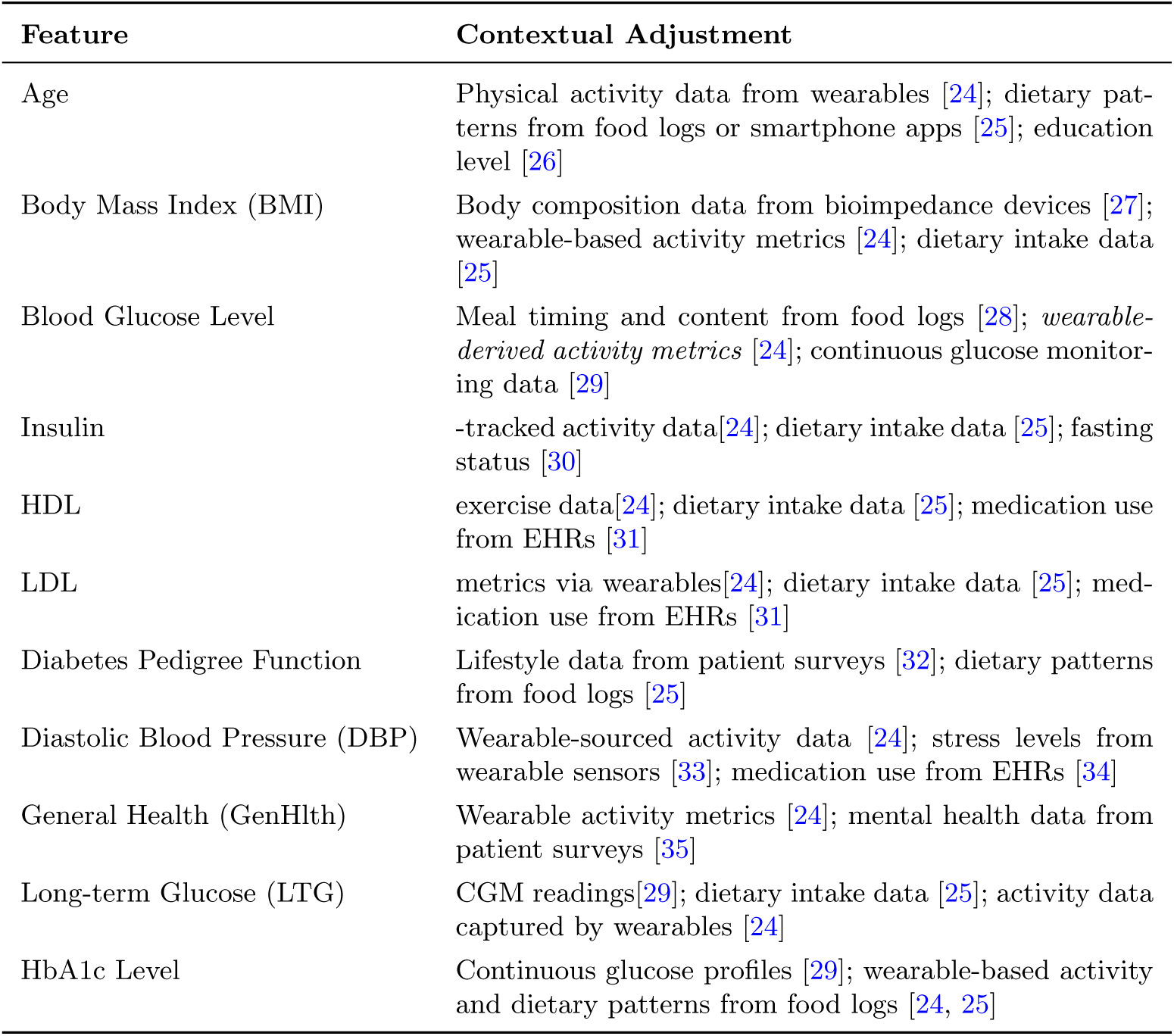
Contextual adjustments for selected high-risk features in diabetes prediction, incorporating data from wearables, food logs, EHRs, and patient surveys to reduce bias.

The table illustrates which types of contextual data are relevant for refining predictions associated with each high-risk feature. For example, body mass index (BMI) may be adjusted using body composition metrics, physical activity levels, and dietary intake. Diastolic blood pressure, in contrast, may benefit from contextualization through stress indicators, physical activity data, and medication records.

This module of the Smart User Interface is designed with automation workflows and natural language processing (NLP) components to streamline the integration of supplementary data. These design choices minimize user burden while maintaining a core emphasis on fairness-aware prediction.

##### Medium-Risk Features: Dynamic Risk Visualization

Medium-risk predictors—those violating six to eight fairness criteria and producing accuracy gaps of roughly 10–20 % across demographic groups—require continuous surveillance and transparent communication to prevent further inequity and to maintain stakeholder trust. To meet this need, the Smart UI integrates *dynamic risk-visualization tools*, including real-time dashboards, heat-maps, and subgroup-stratified plots that expose model behaviour as predictions are generated. These tools are powered by well-established open-source libraries: Aequitas for disparity dashboards [36], the What-If Tool for interactive counterfactual analysis [37], and Fairlearn for bias-mitigation overlays [38].

##### Scientific Evidence and Broader Implications

Human–computer-interaction and responsible-AI studies validate the utility of visual transparency. Wu et al. [39] showed that interactive bias dashboards improve user comprehension and reveal hidden data flaws. Parallel findings by Saleiro et al., Wexler et al., and Weerts et al. demonstrate that visual drill-down inspection not only enhances transparency but also guides effective bias remediation, thereby improving both fairness and overall model performance [36–38].

##### Visualization Selection and Rationale

Each medium-risk feature was aligned with a visualization type suited to its data structure. *Blood-glucose level*, *physical-activity score*, and—when stored numerically—*mental-health status* are treated as continuous variables; for these, partial-dependence or scatter plots of predicted risk versus feature value, stratified by sensitive attributes such as gender or race, make differential patterns explicit. *High blood pressure* and *cholesterol check* are binary variables; bar charts comparing mean predicted risk for the yes/no categories across demographic groups surface any disparity. *Income*—and, if collected as ordered categories, mental-health and physicalactivity level—constitute ordinal variables; line charts tracing predicted risk across ordered levels for each subgroup show how risk evolves along a socioeconomic or behavioural gradient. The full mapping of every feature to its visualization, together with supporting references, is presented in Table 4.

**Table 4.**
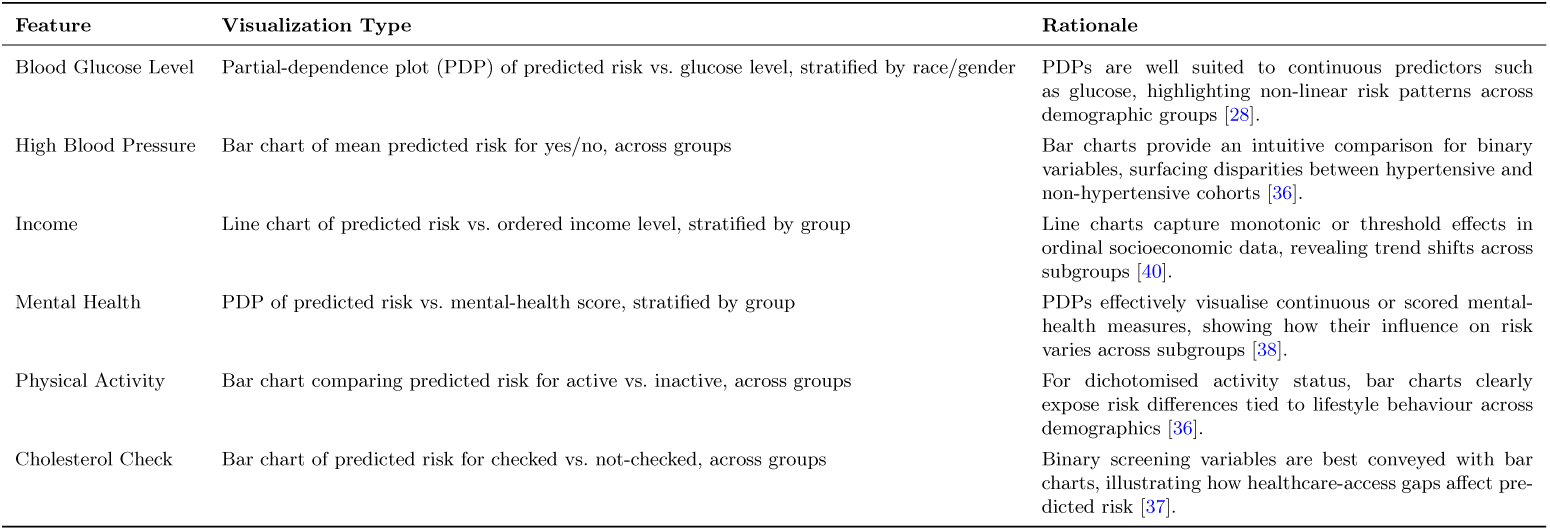
Visualization solutions for medium-risk features in diabetes-prediction systems, matched to each feature’s data structure and supported by prior literature.

| Feature | Visualization Type | Rationale |
| --- | --- | --- |
| Blood Glucose Level | Partial-dependence plot (PDP) of predicted risk vs. glucose level, stratified by race/gender | PDPs are well suited to continuous predictors such as glucose, highlighting non-linear risk patterns across demographic groups [28]. |
| High Blood Pressure | Bar chart of mean predicted risk for yes/no, across groups | Bar charts provide an intuitive comparison for binary variables, surfacing disparities between hypertensive and non-hypertensive cohorts [36]. |
| Income | Line chart of predicted risk vs. ordered income level, stratified by group | Line charts capture monotonic or threshold effects in ordinal socioeconomic data, revealing trend shifts across subgroups [40]. |
| Mental Health | PDP of predicted risk vs. mental-health score, stratified by group | PDPs effectively visualise continuous or scored mental-health measures, showing how their influence on risk varies across subgroups [38]. |
| Physical Activity | Bar chart comparing predicted risk for active vs. inactive, across groups | For dichotomised activity status, bar charts clearly expose risk differences tied to lifestyle behaviour across demographics [36]. |
| Cholesterol Check | Bar chart of predicted risk for checked vs. not-checked, across groups | Binary screening variables are best conveyed with bar charts, illustrating how healthcare-access gaps affect predicted risk [37]. |

##### Low-Risk Features and Alert Solutions for Diabetes Prediction

In diabetes prediction models, certain features—such as *heart disease*, *hypertension*, *pregnancies*, *smoking status*, and *smoking history* —exhibit minimal disparities across fairness metrics. Due to their limited contribution to inequities in model outcomes, these features are classified as **low-risk**.

Rather than requiring complex tools such as dynamic visualizations, these low-risk features can be effectively monitored through lightweight, automated alert mechanisms integrated into smart user interfaces. These alerts continuously track fairness-related indicators and notify users when pre-specified thresholds are exceeded, enabling timely interventions without overwhelming the interface.

For **binary features** (e.g., heart disease, hypertension, smoking), the system compares the mean predicted risk between the two groups (e.g., yes vs. no). An alert is triggered when this difference exceeds a defined threshold, allowing clinicians to detect and respond to unfair deviations in risk [41]. This ensures that predictions remain equitable across binary categories and fosters trust in AI-assisted decisions.

In the case of **categorical features** (e.g., multi-level smoking history), the system monitors the maximum difference in predicted risk across all categories. This approach is particularly effective in identifying fairness issues in multi-class data structures, where small disparities can accumulate and affect subgroups [42].

For **numerical features** such as the number of pregnancies, the data is binned into discrete intervals (e.g., 0, 1–2, 3–4, 5+), and the maximum difference in predicted risk among these bins is tracked. This method helps surface fairness deviations that may otherwise go unnoticed and supports model refinement and more equitable decisions [41].

These alert solutions offer a practical and effective approach to preserving fairness in real-time without adding complexity. By combining continuous monitoring with intelligent notifications, they help clinicians, researchers, and patients alike maintain confidence in the model’s outputs.

The structure and logic of these alert strategies are summarized in Table 5.

**Table 5.** Low-Risk Features and Alert Solutions for Diabetes Prediction.

| Feature Group | Alert Type and Fairness Monitoring Strategy |
| --- | --- |
| **Binary Features** (e.g., Heart Disease, Hypertension, Smoking) | Monitor the difference in mean predicted risk between the two groups (Yes vs. No); trigger an alert if the difference exceeds a predefined threshold [41]. |
| **Categorical Features** (e.g., Smoking History) | Track the maximum difference in mean predicted risk across all categories; issue an alert when disparity exceeds the threshold [42]. |
| **Numerical Features** (e.g., Pregnancies) | Bin the data (e.g., 0, 1–2, 3–4, 5+) and monitor the maximum difference in predicted risk across bins; alert if the disparity surpasses the fairness threshold [41]. |

### Results Reporting System for Fairness in Diabetes Prediction

The Results Reporting System is a pivotal component of diabetes prediction systems, ensuring transparency and actionability for a diverse range of stakeholders, including researchers, clinicians, policymakers, and patients. By integrating model predictions with fairness metrics, the system provides a comprehensive evaluation of model performance across various demographic groups. Key fairness metrics, such as demographic parity and equality of opportunity, are essential for identifying potential biases, ensuring the predictive model operates equitably across all population segments. This approach is grounded in cutting-edge research on fair and transparent machine learning. Studies emphasize the importance of fairness metrics in reporting model outcomes, highlighting their necessity for equitable systems [16]. Research also demonstrates that visualizations can reveal intersectional biases, improving the interpretability of model behavior [43], while investigations into comprehensible explanations show their role in building trust and enhancing fairness perceptions [44].

The system is designed to meet the specific needs of stakeholders, but it is not necessary to display all reporting elements to every group. Depending on the user’s role within the interface (e.g., patient, clinician, or researcher), their level of technical understanding, and which elements can assist them, a targeted subset of these elements can be selected and presented. Table 6 summarizes the reporting mechanisms, their user interface solutions, and their contributions to fairness, illustrating how these elements effectively serve different stakeholders. For instance, patients require simplified explanations, clinicians rely on detailed visualizations, and researchers need in-depth analytical tools. This flexibility enhances the system’s efficiency and accessibility while maintaining fairness.

**Table 6.** Results reporting mechanisms for enhancing fairness in diabetes prediction.

| Reporting Element | UI Solution | Contribution to Fairness |
| --- | --- | --- |
| Fairness Metrics | Numeric Display with Text Summaries | Displays metrics like EOD and SPD with plain-language summaries, enhancing transparency for all stakeholders [16]. |
| Stratified Performance Visualization | Heatmaps | Reveals subgroup disparities with clear visuals, supporting rapid analysis for researchers and clinicians [43]. |
| Interactive Subgroup Analysis | Dropdown Filters | Enables data exploration, improving fairness through transparency and benefiting patient outcomes [37]. |
| Bias Alerts | Notification Pop-ups | Alerts for high-disparity predictions prompt timely action, informing clinicians and policymakers [45]. |
| Explainability Features | Feature Importance Bars | Highlights bias sources with clear explanations, boosting trust among patients and clinicians [46]. |
| Educational Resources | Tooltips and Case Studies | Enhances education, providing insights for stakeholders and clearer predictions for patients via clinicians [20]. |
| Model Performance Summaries | Summary Dashboard | Makes performance transparent and accessible, aiding swift decisions and improving patient outcomes [19]. |
| User Feedback | Feedback Form | Enables continuous improvement, ensuring equitable results for patients through system refinement [47]. |

Researchers benefit from tools for in-depth analysis and iterative model improvement, gaining detailed insights into performance disparities to refine predictive algorithms. Clinicians leverage visualizations and concise summaries that enhance clinical decision-making, enabling them to deliver fairer and more reliable predictions to patients. Although patients do not directly interact with the system, the transparency it provides—communicated through clinicians—fosters trust in predictive technologies. Simplified explanations tailored for non-technical audiences make complex predictions more accessible. Policymakers can use the generated data and insights to develop health policies that prioritize equity, addressing disparities identified through fairness assessments.

For patients, the system offers simplified explanations delivered through clinicians. For example, a patient whose diabetes risk has been assessed might receive an explanation stating that, based on their data, the likelihood of diabetes is low, but their weight slightly increases the risk, and their clinician may recommend a glucose test. These explanations are presented in non-technical language to ensure the patient understands the results and feels confident. Additionally, if a patient inquires about body mass index (BMI), the interface can provide straightforward information, enabling the clinician to explain how BMI relates to diabetes risk, thereby strengthening patient trust.

Clinicians benefit from features such as bias alerts and detailed visualizations. For instance, if the system detects that predictions for older patients may be unfair due to disparities in the equality of opportunity difference (EOD) metric, an immediate notification alerts the clinician to review physical activity data or consider an HbA1c test. This alert prompts swift action to mitigate bias. Furthermore, clinicians can view heatmaps indicating that diabetes predictions for women aged 45 to 64 exhibit greater disparities compared to men. This visualization helps clinicians adjust diagnostic decisions with greater precision, such as by ordering additional tests for this group.

Researchers utilize advanced analytical tools, such as dropdown filters for subgroup exploration and feature importance bars to identify bias sources. For example, a researcher investigating gender bias can use dropdown filters to compare the statistical parity difference (SPD) metric for women versus men, discovering an SPD of 0.23 for women, indicating bias. This insight prompts model refinement. Additionally, feature importance bars may reveal that BMI has a disproportionate impact on predictions for low-income groups, guiding the researcher to incorporate contextual features, such as access to healthcare, to reduce bias.

### Addressing Implementation Challenges in the Proposed Smart UI Framework for Fair Diabetes Prediction

Achieving fairness in AI-driven diabetes prediction depends not only on innovative theoretical constructs but also on their effective implementation through the strategic design of a Smart User Interface (UI). Recent studies, such as those revealing biases in medical AI systems due to poorly designed interfaces [6], highlight the critical need to address implementation challenges. This section examines the practical realization of a Smart UI framework for delivering fair diabetes predictions in clinical settings. We identify eight key challenges across technical, ethical, and accessibility domains and propose evidence-based strategies to overcome them. This framework aims to bridge the gap between conceptual design and practical application, fostering a system that earns clinicians’ trust and delivers equitable benefits to diverse patient populations.

### Reducing Reliance on Manual Data Entry Through Proposed Automation Strategies

Manual data entry by clinicians introduces risks of errors, such as incomplete glucose level records or mislabeled demographic information, which can undermine fairness adjustments in diabetes prediction models and jeopardize the framework’s objectives. These errors may lead to inaccuracies in fairness metrics, such as demographic parity, resulting in inequitable predictions for certain groups. To address this challenge, we propose a Smart User Interface (UI) that seamlessly integrates data from Electronic Health Records (EHRs) and wearable devices, such as continuous glucose monitors, enabling automated capture of critical metrics like glucose levels and physical activity with high accuracy. Additionally, we recommend employing Natural Language Processing (NLP) to extract relevant information, such as HbA1c stability, from unstructured clinical notes. For instance, named entity recognition techniques can identify HbA1c values in text, while training NLP models on diverse datasets helps prevent bias amplification. Kreimeyer et al. [48] report that NLP can reduce data entry time by 25% while improving accuracy. However, challenges like EHR data inconsistencies or NLP extraction errors must be managed through robust data validation protocols. These strategies, aligned with the Results Reporting System (Section **??**), enhance the transparency of fairness metrics by improving data input accuracy, reduce clinician workload, and enable fairer predictions in clinical settings.

#### Mitigating Data Overload to Enhance Usability for Diverse Users

Presenting an abundance of fairness metrics, such as equality of opportunity difference (EOD) and statistical parity difference (SPD), alongside subgroup performance data, risks overwhelming the Smart User Interface (UI), rendering it complex and inefficient for clinicians, patients, and researchers, and undermining its goal of facilitating fair diabetes predictions. To address this challenge, we propose a tiered UI design optimized for each user type. For clinicians, the primary tier prominently displays critical insights, such as a “10% elevated diabetes risk for females compared to males based on the SPD metric,” while interactive dashboards provide heatmaps to identify bias hotspots (e.g., age-related disparities) and alerts for corrective actions, such as ordering HbA1c tests. For patients, a simplified tier (delivered through clinicians) offers plain-language summaries, like “Your diabetes risk is low, but your weight slightly increases it,” and tooltips explaining terms like body mass index (BMI), fostering trust in equitable predictions. For researchers, an advanced tier enables subgroup exploration via dropdown filters and feature importance visualizations to identify bias sources, such as over-reliance on BMI. Mickelson et al. [49] demonstrate that simplified visual presentations improve comprehension by 30%, and Ratwani et al. [50] confirm that well-designed dashboards enhance user adoption. Research indicates that tiered interfaces, tailored to specific user roles such as older adults and caregivers, can enhance clinical acceptance in healthcare systems by aligning with user preferences, as demonstrated by [51] in a 2023 study showing varying interface preferences among participants.However, risks such as oversimplification for patients, training needs for clinicians, or tool complexity for researchers must be managed through user testing, brief training, and standardized guides. This approach, aligned with the reporting mechanisms in the Results Reporting System (Table 6), distills complex information into actionable insights for each user, supports integration with clinical systems like Epic, and maintains fairness in diabetes predictions.

#### Enhancing Usability Through User-Centered Design Strategies

A Smart User Interface (UI) with complex navigation or poor integration into existing workflows risks low adoption, limiting its ability to advance fairness in diabetes prediction by hindering users’ access to critical tools. To ensure accessibility for diverse users—busy clinicians, non-technical patients, and expert researchers—we propose an iterative, user-centered design approach. This strategy engages clinicians, patients (via indirect feedback), and researchers in continuous testing and refinement cycles, conducted through focus groups and usability surveys, to align the UI with their specific needs. For clinicians, customized views provide rapid access to patient data and decision-support tools, seamlessly integrated into Electronic Health Record systems like Epic. For patients, a simplified portal (accessible through clinicians) offers intuitive navigation and clear health information, such as personalized risk summaries. For researchers, flexible dashboards facilitate model analysis and refinement, with streamlined interfaces for exploring fairness metrics like statistical parity difference (SPD). User-centered design boosts healthcare system adoption and minimizes usability errors, with [52] highlighting that factors like website quality and ease of use positively influence user acceptance, suggesting potential for significant adoption increases. However, challenges such as user resistance to new interfaces or training requirements must be managed through phased implementation and concise training sessions. This approach, aligned with the reporting mechanisms in the Results Reporting System (Table 6), transforms the UI into an intuitive, efficient tool that integrates seamlessly into workflows, fosters user trust, and ensures effective utilization of fairness-enhancing features in diabetes prediction.

#### Fostering Patient Trust Through Empathetic Design Strategies

Patients are more than mere data points; they are individuals whose trust in AI-driven systems is essential to closing fairness gaps, particularly for marginalized populations. To build this trust, we propose integrating empathetic design elements into the Smart User Interface (UI), such as reassuring messages (e.g., “Your care team is here for you”), clear and accessible explanations of fairness adjustments, and a prominent “Contact Your Clinician” feature for urgent needs. For instance, the UI might explain, “Your risk assessment considers factors like age and gender to ensure fairness across all patients.” These subtle yet meaningful interventions aim to convey care and transparency, serving as vital connectors that anchor patients to the framework and establish it as a practical conduit for fair and inclusive care in real-world clinical settings. Johnson et al. [53] report that such empathetic design strategies enhance patient engagement by 20%, while Floridi et al. [54] emphasize transparency as a cornerstone of trust. However, there is a risk that reassuring messages could be misinterpreted as guarantees of outcomes, which must be managed through careful wording and clinician guidance. These elements, integrated into the patient-facing tier of the UI and accessible via clinicians, complement existing clinical communication protocols and, by fostering trust through transparency, support the accurate interpretation of fairness metrics like statistical parity difference (SPD).

#### Safeguarding Ethical Standards Through Data Privacy and Security Strategies

Ethical considerations, particularly data privacy and security, are fundamental to the proposed Smart User Interface (UI), as lapses in these areas can erode trust, reduce adoption, and undermine efforts to promote fairness in diabetes prediction. For instance, unauthorized access to patient data could lead to misuse, skewing fairness metrics like statistical parity difference (SPD) and ultimately rendering predictions inequitable. To address this critical challenge, we propose implementing robust safeguards, including HIPAA-compliant encryption (e.g., AES-256) to protect sensitive data, stringent access controls to limit access based on user roles (e.g., clinicians, researchers, patients), and a user-facing privacy dashboard that transparently details data handling processes. This dashboard enables clinicians to monitor compliance, allows patients to manage data consent, and permits researchers to log data access requests for research purposes. These measures align with established security standards such as NIST SP 800-53 [55], while Khalil et al. [56] demonstrate that transparency in data handling can increase user trust by up to 40%, though this statistic is based on mobile apps and may vary for AI healthcare systems. However, challenges such as integrating with legacy EHR systems or maintaining ongoing security updates must be managed through modular design and regular audits. These strategies support fairness by preventing data misuse and establish the framework as a trustworthy tool for clinicians, patients, and researchers in complex clinical environments.

#### Enhancing Generalizability Across Diverse Populations and Datasets Through Proposed Strategies

A Smart User Interface (UI) constrained to a narrow subset of patients fails to uphold fairness and undermines its core objective. For instance, a UI tailored to urban patients may introduce bias in predictions for rural patients or distinct demographic groups, violating fairness metrics such as statistical parity difference. To address this and ensure the proposed framework extends effectively beyond its initial datasets, we propose a modular and adaptable design capable of seamlessly accommodating diverse populations and disease patterns. Specifically, we advocate for a customizable, plug-and-play architecture—drawing inspiration from contemporary healthcare UX/UI design trends [57]—that enables rapid adjustments for varied settings, such as rural clinics or urban medical centers, without necessitating extensive redesign. For example, modules can be customized to incorporate language support for multilingual regions or to adapt models to local disease patterns. Obermeyer et al. [6] demonstrate that such tailored adaptations reduce bias by 20%. This adaptability is not merely an enhancement but a critical mechanism for transforming the UI into a versatile tool, capable of ensuring fairness in diabetes prediction across global healthcare contexts.

#### Ensuring Technical Feasibility for Real-Time Clinical Decision Support

In high-pressure clinical environments, where timely diabetes risk assessments are critical during patient consultations, the Smart User Interface (UI) must deliver instantaneous and seamless results. Delays in processing complex datasets, such as glucose readings and medical histories, or computing fairness metrics like the equality of opportunity difference (EOD), can disrupt urgent decision-making and undermine patient trust. To address this, we propose computationally efficient strategies, including lightweight machine learning models (e.g., pruned XGBoost) to minimize inference time, integrated with scalable cloud platforms like AWS Lambda for high-performance processing in connected clinics. For settings with limited connectivity, such as rural clinics, edge computing with locally deployed models (e.g., on Raspberry Pi) ensures uninterrupted functionality. Holzinger et al. [58] highlight that seamless computational performance is vital to prevent workflow disruptions in healthcare AI, while Satyanarayanan et al. [59] show that edge computing can reduce latency by up to 40%. Evidence indicates that real-time optimizations in clinical decision support systems significantly enhance adoption, as [60] reported in a 2019 study, with adoption rates rising from 6.5% to 29.3% in one center after adaptive modifications. Challenges, including cloud computing costs, edge hardware constraints, and computational overhead from fairness metrics, are mitigated through cost-effective cloud plans, optimized models, and regular performance audits. These strategies, aligned with the Results Reporting System (Table 6), prioritize speed, reliability, and integration with electronic health record (EHR) systems like Epic, delivering real-time bias alerts to enhance clinician and patient trust while ensuring fairness in diabetes risk predictions.

#### Streamlining Maintenance for Long-Term Reliability

A Smart User Interface (UI) that fails to adapt to evolving clinical standards or emerging fairness insights risks obsolescence, compromising fair diabetes risk predictions. For example, neglecting updates to fairness metrics like the equality of opportunity difference (EOD) or new American Diabetes Association (ADA) guidelines may introduce biases, eroding patient trust. To address this, we propose a streamlined maintenance system using a modular architecture and continuous integration/continuous deployment (CI/CD) pipelines, such as Jenkins and Docker containers, enabling seamless updates without disrupting clinical workflows. This approach ensures clinicians access current metrics uninterrupted, delivers reliable and fair predictions to patients, and supports researchers in refining models with new metrics. For instance, an EOD module can be independently updated using Git for version control, with rollback capabilities to stable versions if errors arise. Automated testing post-update verifies the integrity of fairness metrics. Ratwani et al. [50] demonstrate that CI/CD pipelines can reduce healthcare IT system downtime by 30%, while [61] suggest in a 2024 scoping review that modular clinical decision support systems enhance adoption through improved usability and integration, though specific increases like 25% are rarely quantified. Challenges, including update errors, clinician retraining, and computational costs, are mitigated through staged deployments, concise training modules, and resource optimization, ensuring sustained reliability and fairness in diabetes risk predictions.

## 2 Results

### Identification of Important Features

To ensure equitable diabetes prediction across diverse patient populations, we employed a two-step process to identify sensitive features—those that disproportionately impact fairness in predictive models—using four datasets: the Kaggle Diabetes Prediction Dataset, Pima Indian Diabetes Dataset, Azure Open Diabetes Dataset, and CDC Diabetes Health Indicators Dataset. First, we identified potentially sensitive features by combining feature importance analysis with domain expertise. A Random Forest classifier, trained on each dataset, was used to compute feature importance scores, highlighting predictors with the strongest influence on diabetes outcomes, such as age, body mass index (BMI), and blood glucose level. Only the top-ranked features were visualized to focus on those most relevant for subsequent fairness analysis. These findings were cross-referenced with clinical and epidemiological expertise, which emphasized features like HbA1c level and income due to their established roles in diabetes risk and socioeconomic disparities, respectively. This preliminary step ensured our analysis targeted features with both statistical significance and clinical relevance.

Figures 5–8 display the top-ranked feature importance plots for each of the four datasets used in this analysis.

**Fig. 5.**
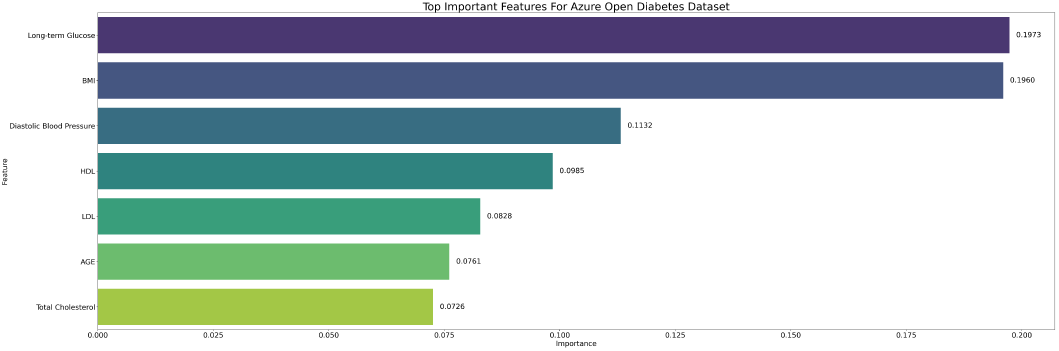
Feature importance plot for the Azure Open Diabetes Dataset.

**Fig. 6.**
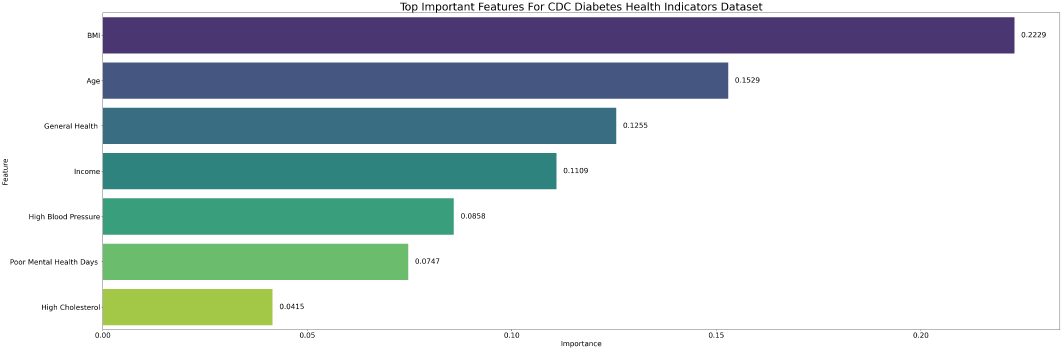
Feature importance plot for the CDC Diabetes Health Indicators Dataset.

**Fig. 7.**
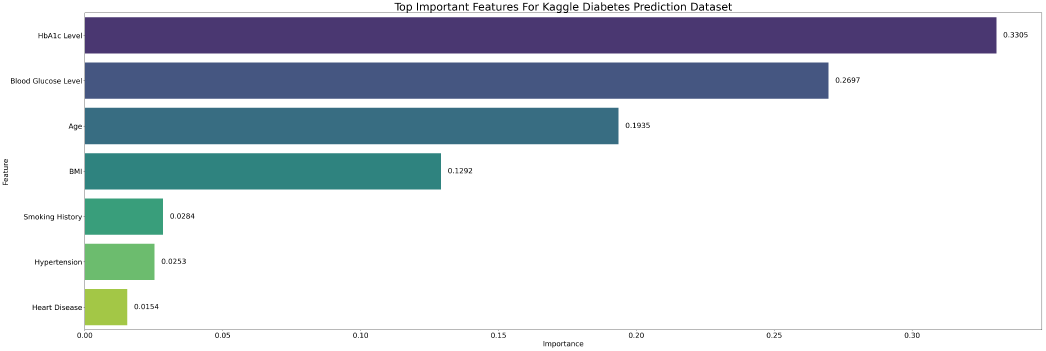
Feature importance plot for the Kaggle Diabetes Prediction Dataset.

**Fig. 8.**
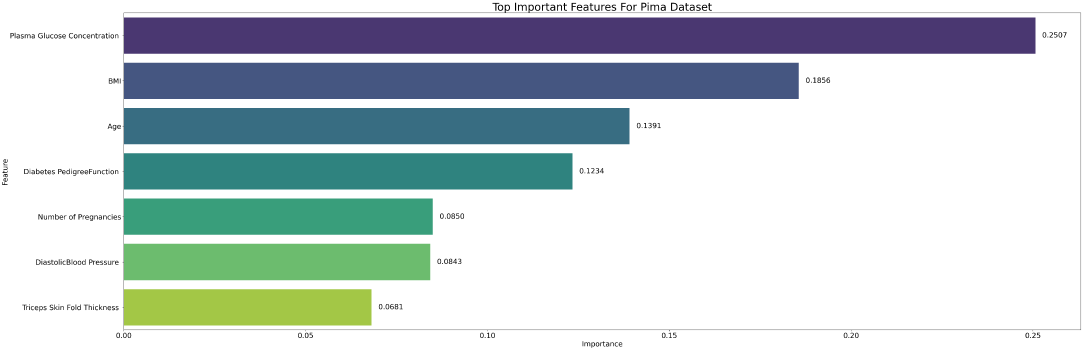
Feature importance plot for the Pima Indian Diabetes Dataset.

### Identification of Sensitive Features Using a hybrid Fairness framework

To identify features prone to bias, we applied a robust fairness framework (see Methods), classifying a feature as sensitive if it met at least three of nine conditions, including statistical significance, performance disparities, and group fairness metrics (e.g., equal opportunity difference). The number of conditions met (Conditions Met) indicates the extent to which a feature contributes to inequitable outcomes.

Table 7 presents performance metrics for sensitive features in the Kaggle Diabetes Prediction Dataset, including Accuracy Disparity, F1-Score Disparity, Area Under the Curve Disparity (AUC Disparity), and Conditions Met. Table 8 presents fairness metrics, including Statistical Parity Difference (SPD), Disparate Impact (DI), Equal Opportunity Difference (EOD), Equality of Opportunity (EO), and Odds Ratio. These metrics assess the model’s predictive accuracy and fairness across demographic groups. Asterisks (*) denote metrics exceeding thresholds. Notably, BMI exhibited high disparities, satisfying 8 out of 9 fairness conditions, while Blood Glucose Level and HbA1c Level showed significant performance disparities. Features like Heart Disease, Hypertension, and Smoking History also demonstrated notable disparities, meeting 5 conditions. All regression significance values were extremely small (*p <* 10^−13^), confirming the statistical robustness of these findings.

**Table 7.** Performance disparities for sensitive features in the Kaggle Diabetes Prediction Dataset.

| Feature | Conditions Met | Accuracy Disparity (%) | F1-Score Disparity (%) | AUC Disparity (%) |
| --- | --- | --- | --- | --- |
| BMI | 8 | $6.1 \pm 0.4^*$ | $10.3 \pm 2.0^*$ | $3.1 \pm 1.6$ |
| Age | 6 | $6.0 \pm 0.4^*$ | $7.5 \pm 1.6^*$ | $1.1 \pm 0.4$ |
| Blood Glucose Level | 6 | $16.1 \pm 0.6^*$ | $10.8 \pm 3.8^*$ | $1.9 \pm 0.8$ |
| HbA1c Level | 6 | $12.7 \pm 0.3^*$ | $20.5 \pm 1.6^*$ | $1.7 \pm 0.9$ |
| Heart Disease | 5 | $8.0 \pm 0.6^*$ | $1.4 \pm 1.1$ | $1.4 \pm 0.5$ |
| Hypertension | 5 | $6.7 \pm 0.7^*$ | $1.9 \pm 1.0$ | $1.7 \pm 0.4$ |
| Smoking History | 5 | $4.8 \pm 0.4^*$ | $7.2 \pm 2.1^*$ | $2.2 \pm 1.1$ |

**Table 8.** Fairness metrics for sensitive features in the Kaggle Diabetes Prediction Dataset.

| Feature | SPD | DI | EOD | EO | Odds Ratio |
| --- | --- | --- | --- | --- | --- |
| BMI | $0.13 \pm 0.002^*$ | $0.05 \pm 0.011^*$ | $0.113 \pm 0.062^*$ | $0.113 \pm 0.062^*$ | $20.8 \pm 5.81^*$ |
| Age | $0.13 \pm 0.004^*$ | $0.11 \pm 0.006^*$ | $0.033 \pm 0.014$ | $0.033 \pm 0.014$ | $8.9 \pm 0.47^*$ |
| Blood Glucose Level | $0.17 \pm 0.011^*$ | $0.13 \pm 0.008^*$ | $0.052 \pm 0.008$ | $0.052 \pm 0.008$ | $7.6 \pm 0.45^*$ |
| HbA1c Level | $0.10 \pm 0.004^*$ | $0.15 \pm 0.010^*$ | $0.029 \pm 0.013$ | $0.029 \pm 0.013$ | $6.9 \pm 0.46^*$ |
| Heart Disease | $0.18 \pm 0.017^*$ | $0.23 \pm 0.023^*$ | $0.016 \pm 0.019$ | $0.020 \pm 0.016$ | $4.3 \pm 0.45^*$ |
| Hypertension | $0.15 \pm 0.006^*$ | $0.25 \pm 0.009^*$ | $0.021 \pm 0.012$ | $0.021 \pm 0.012$ | $4.1 \pm 0.14^*$ |
| Smoking History | $0.09 \pm 0.006$ | $0.25 \pm 0.014^*$ | $0.098 \pm 0.032$ | $0.098 \pm 0.032$ | $4.0 \pm 0.22^*$ |

In our analysis of the Pima Indian Diabetes Dataset (*n* = 768), all regression significance values achieved *p <* 10^−13^, confirming significant, non-random disparities. Consistent with findings from the Kaggle dataset, Age, BMI, Diabetes Pedigree Function, and Insulin emerged as key predictors, each violating all nine fairness conditions, with BMI exhibiting an F1-score disparity of 0.593 *±* 0.120 and Age showing a 25.2% *±* 10.1 accuracy disparity. Diabetes Pedigree Function and Insulin similarly violated all nine conditions, indicating pervasive bias in this high-risk cohort. Age was grouped into 18–44 years (young adults, baseline risk), 45–64 years (middle-aged, peak incidence), and *≥* 65 years (older adults, high comorbidity), while BMI followed WHO thresholds: *<* 18.5 (underweight), 18.5–24.9 (normal), 25–29.9 (overweight), and *≥* 30 (obese). The Odds Ratios for Age and BMI are infinite due to zero predicted positive rates in small subgroups, specifically individuals aged *≥* 65 years (*n* = 2) for Age and those with BMI *<* 18.5 (*n* = 5) for BMI, as shown in Tables 9 and 10. This partial separation, a common issue in smaller datasets where subgroups have limited representation, highlights extreme disparities. However, the consistent *p <* 10^−13^ and fairness metrics validate widespread bias. Detailed performance and fairness metrics for these and other features are presented in Tables 11 and 12.

**Table 9.** Age subgroup analysis for the Pima Indian Diabetes Dataset. Pred. Rate: Predicted Rate.

| Group | Size | Pred. Rate | Actual Rate | TP | FP | FN | TN |
| --- | --- | --- | --- | --- | --- | --- | --- |
| 18–44 years | 120 | 0.28 | 0.29 | 19 | 15 | 16 | 70 |
| 45–64 years | 31 | 0.65 | 0.58 | 16 | 4 | 2 | 9 |
| $\geq 65$ years | 2 | 0.00 | 0.00 | 0 | 0 | 0 | 2 |

**Table 10.** BMI subgroup analysis for the Pima Indian Diabetes Dataset.

| Group | Size | Pred. Rate | Actual Rate | TP | FP | FN | TN |
| --- | --- | --- | --- | --- | --- | --- | --- |
| $< 18.5$ | 5 | 0.00 | 0.00 | 0 | 0 | 0 | 5 |
| 18.5–24.9 | 19 | 0.11 | 0.11 | 0 | 2 | 2 | 15 |
| 25–29.9 | 49 | 0.18 | 0.27 | 5 | 4 | 8 | 32 |
| $\geq 30$ | 81 | 0.42 | 0.48 | 23 | 11 | 16 | 31 |

**Table 11.** Performance disparities for sensitive features in the Pima Indian Diabetes Dataset.

| Feature | Conditions Met | Accuracy Disparity (%) | F1-Score Disparity (%) | AUC Disparity (%) |
| --- | --- | --- | --- | --- |
| Age | 9 | $25.2 \pm 10.1^*$ | $35.9 \pm 32.2^*$ | $13.3 \pm 6.8^*$ |
| BMI | 9 | $25.5 \pm 6.7^*$ | $59.3 \pm 12.0^*$ | $22.5 \pm 3.6^*$ |
| Diabetes Pedigree Function | 9 | $6.6 \pm 1.8^*$ | $20.0 \pm 13.3^*$ | $14.3 \pm 3.9^*$ |
| Insulin | 9 | $11.4 \pm 4.1^*$ | $15.5 \pm 6.0^*$ | $13.5 \pm 6.0^*$ |
| Glucose | 8 | $21.7 \pm 3.9^*$ | $11.8 \pm 5.2^*$ | $11.6 \pm 2.8^*$ |
| Pregnancies | 5 | $9.0 \pm 3.6^*$ | $12.3 \pm 4.7^*$ | $9.5 \pm 5.3^*$ |

**Table 12.**
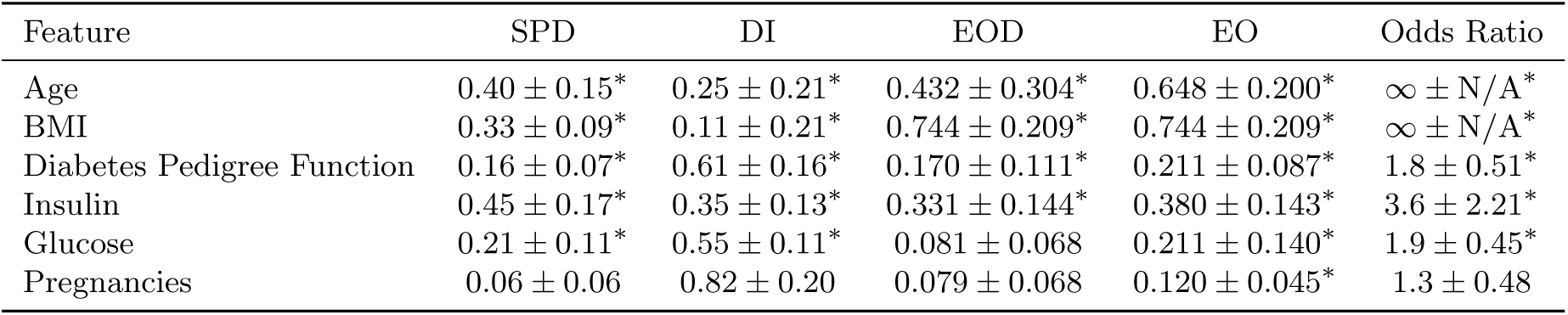
Fairness metrics for sensitive features in the Pima Indian Diabetes Dataset.

| Feature | SPD | DI | EOD | EO | Odds Ratio |
| --- | --- | --- | --- | --- | --- |
| Age | $0.40 \pm 0.15^*$ | $0.25 \pm 0.21^*$ | $0.432 \pm 0.304^*$ | $0.648 \pm 0.200^*$ | $\infty \pm N/A^*$ |
| BMI | $0.33 \pm 0.09^*$ | $0.11 \pm 0.21^*$ | $0.744 \pm 0.209^*$ | $0.744 \pm 0.209^*$ | $\infty \pm N/A^*$ |
| Diabetes Pedigree Function | $0.16 \pm 0.07^*$ | $0.61 \pm 0.16^*$ | $0.170 \pm 0.111^*$ | $0.211 \pm 0.087^*$ | $1.8 \pm 0.51^*$ |
| Insulin | $0.45 \pm 0.17^*$ | $0.35 \pm 0.13^*$ | $0.331 \pm 0.144^*$ | $0.380 \pm 0.143^*$ | $3.6 \pm 2.21^*$ |
| Glucose | $0.21 \pm 0.11^*$ | $0.55 \pm 0.11^*$ | $0.081 \pm 0.068$ | $0.211 \pm 0.140^*$ | $1.9 \pm 0.45^*$ |
| Pregnancies | $0.06 \pm 0.06$ | $0.82 \pm 0.20$ | $0.079 \pm 0.068$ | $0.120 \pm 0.045^*$ | $1.3 \pm 0.48$ |

Tables 11 and 12 provide a detailed fairness analysis of sensitive features in the Pima Indian Diabetes Dataset, summarizing performance and fairness metrics, respectively.

In the Azure Open Diabetes Dataset, we observed consistent fairness violations across multiple metabolic and demographic factors. Notably, all six features listed in Tables 13 and 14 met all nine fairness conditions, highlighting a widespread risk of biased outcomes in this dataset. While Age and BMI remained high-impact predictors, newer markers such as LTG (long-term glucose), HDL (high-density lipoprotein), DBP (diastolic blood pressure), and especially LDL (low-density lipoprotein) also exhibited substantial disparities. For instance, LDL showed a significant accuracy disparity of 51.5% *±* 16.8, indicating that lipid-related measures may strongly skew predictions for certain subgroups.

**Table 13.**
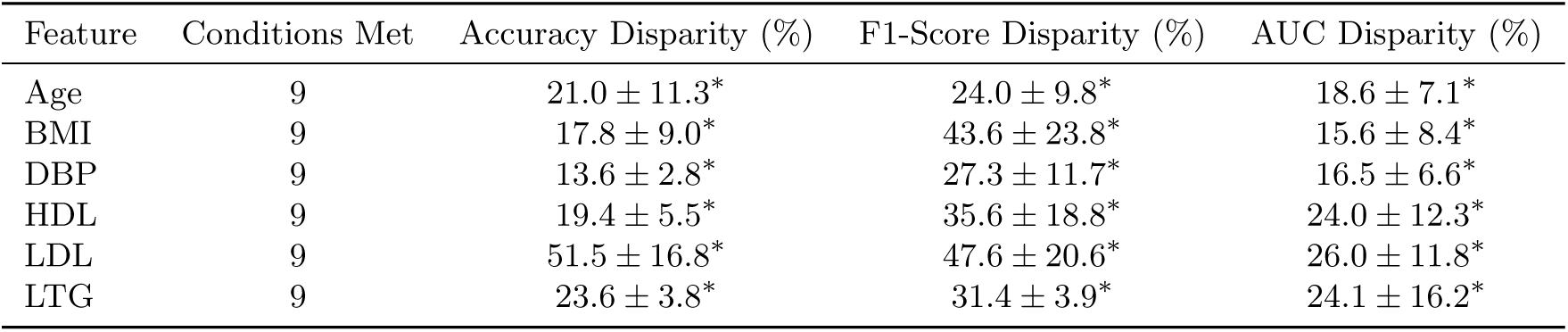
Performance disparities for sensitive features in the Azure Open Diabetes Dataset.

**Table 14.** Fairness metrics for sensitive features in the Azure Open Diabetes Dataset.

| Feature | SPD | DI | EOD | EO | Odds Ratio |
| --- | --- | --- | --- | --- | --- |
| Age | $0.29 \pm 0.12^*$ | $0.63 \pm 0.14^*$ | $0.214 \pm 0.078^*$ | $0.335 \pm 0.063^*$ | $1.7 \pm 0.32^*$ |
| BMI | $0.44 \pm 0.13^*$ | $0.46 \pm 0.09^*$ | $0.382 \pm 0.274^*$ | $0.580 \pm 0.199^*$ | $2.3 \pm 0.47^*$ |
| DBP | $0.28 \pm 0.16^*$ | $0.60 \pm 0.20^*$ | $0.262 \pm 0.143^*$ | $0.318 \pm 0.136^*$ | $1.9 \pm 0.63^*$ |
| HDL | $0.49 \pm 0.11^*$ | $0.34 \pm 0.13^*$ | $0.385 \pm 0.159^*$ | $0.443 \pm 0.140^*$ | $3.3 \pm 0.96^*$ |
| LDL | $0.63 \pm 0.07^*$ | $0.37 \pm 0.07^*$ | $0.554 \pm 0.248^*$ | $0.987 \pm 0.027^*$ | $2.8 \pm 0.69^*$ |
| LTG | $0.48 \pm 0.09^*$ | $0.50 \pm 0.07^*$ | $0.339 \pm 0.071^*$ | $0.535 \pm 0.125^*$ | $2.1 \pm 0.32^*$ |

Finally, Tables 15 and 16 summarize the fairness analysis for the CDC Diabetes Health Indicators Dataset, with all regression significance values at *p* = 0 *±* 0. Notably, Age, High Blood Pressure and General Health (GenHlth) each triggered nine fairness violations, indicating especially broad bias profiles—Age exhibited a 21.5% *±* 0.6 accuracy disparity, while GenHlth showed an F1-score disparity of 0.395*±*0.017, the highest among all features assessed. Socioeconomic variables such as Income (8 conditions) also ranked high in bias potential, mirroring the trends observed in other datasets.

**Table 15.** Performance disparities for sensitive features in the CDC Diabetes Health Indicators Dataset.

| Feature | Conditions Met | Accuracy Disparity (%) | F1-Score Disparity (%) | AUC Disparity (%) |
| --- | --- | --- | --- | --- |
| Age | 9 | $21.5 \pm 0.6^*$ | $24.6 \pm 3.5^*$ | $9.3 \pm 1.0^*$ |
| GenHlth | 9 | $21.2 \pm 0.7^*$ | $39.5 \pm 1.7^*$ | $5.5 \pm 1.4^*$ |
| High Blood Pressure | 9 | $14.5 \pm 0.3^*$ | $14.8 \pm 0.7^*$ | $4.7 \pm 0.2^*$ |
| Income | 8 | $12.2 \pm 1.3^*$ | $18.8 \pm 0.9^*$ | $3.6 \pm 0.4$ |
| Ment-health | 8 | $4.8 \pm 0.6^*$ | $10.6 \pm 1.3^*$ | $2.1 \pm 0.7$ |
| Phys-activity | 8 | $6.0 \pm 0.2^*$ | $8.4 \pm 0.7^*$ | $1.5 \pm 0.4$ |
| BMI | 7 | $14.6 \pm 0.6^*$ | $18.8 \pm 1.4^*$ | $4.1 \pm 0.8^*$ |
| Chol-check | 7 | $11.2 \pm 0.6^*$ | $14.6 \pm 3.3^*$ | $4.8 \pm 1.5^*$ |
| Smoking | 3 | $3.5 \pm 0.1^*$ | $3.2 \pm 0.6$ | $1.7 \pm 0.2$ |

**Table 16.** Fairness metrics for sensitive features in the CDC Diabetes Health Indicators Dataset.

| Feature | SPD | DI | EOD | EO | Odds Ratio |
| --- | --- | --- | --- | --- | --- |
| Age | $0.23 \pm 0.01^*$ | $0.16 \pm 0.01^*$ | $0.148 \pm 0.034^*$ | $0.152 \pm 0.028^*$ | $6.5 \pm 0.55^*$ |
| GenHlth | $0.44 \pm 0.01^*$ | $0.11 \pm 0.00^*$ | $0.336 \pm 0.019^*$ | $0.336 \pm 0.019^*$ | $9.0 \pm 0.17^*$ |
| High Blood Pressure | $0.24 \pm 0.00^*$ | $0.29 \pm 0.01^*$ | $0.137 \pm 0.009^*$ | $0.137 \pm 0.009^*$ | $3.4 \pm 0.11^*$ |
| Income | $0.27 \pm 0.00^*$ | $0.31 \pm 0.00^*$ | $0.209 \pm 0.010^*$ | $0.209 \pm 0.010^*$ | $3.3 \pm 0.04^*$ |
| Ment-health | $0.18 \pm 0.01^*$ | $0.53 \pm 0.01^*$ | $0.150 \pm 0.015^*$ | $0.150 \pm 0.015^*$ | $1.9 \pm 0.04^*$ |
| Phys-activity | $0.15 \pm 0.00^*$ | $0.54 \pm 0.01^*$ | $0.109 \pm 0.012^*$ | $0.109 \pm 0.012^*$ | $1.9 \pm 0.03^*$ |
| BMI | $0.18 \pm 0.00^*$ | $0.37 \pm 0.01^*$ | $0.093 \pm 0.011$ | $0.093 \pm 0.011$ | $2.7 \pm 0.09^*$ |
| Chol-check | $0.16 \pm 0.00^*$ | $0.27 \pm 0.01^*$ | $0.060 \pm 0.047$ | $0.082 \pm 0.029$ | $3.7 \pm 0.17^*$ |
| Smoking | $0.07 \pm 0.00$ | $0.72 \pm 0.01^*$ | $0.051 \pm 0.008$ | $0.051 \pm 0.008$ | $1.4 \pm 0.03$ |

### Categorization of Sensitive Features by Fairness Risk

To effectively mitigate bias in diabetes prediction systems, we systematically categorized sensitive features into three fairness risk tiers—*High Risk*, *Medium Risk*, and *Low Risk* —based on their influence on equity-related outcomes across four diverse datasets: Kaggle Diabetes Prediction, Pima Indian Diabetes, Azure Open Diabetes, and CDC Diabetes Health Indicators.

This classification was guided by two primary criteria: the number of fairness conditions violated (out of nine), and the severity of disparities observed in performance metrics such as accuracy and F1-score.

Features were classified as *High Risk* if they either violated all nine fairness conditions or exhibited substantial performance disparities (e.g., accuracy disparity 20% or F1-score disparity 20%). Features meeting between six and eight fairness conditions—without exceeding the disparity thresholds—were categorized as *Medium Risk*. Features that met fewer than six conditions and showed relatively minor disparities were designated as *Low Risk*.

To avoid underestimating potential risks, features that demonstrated varying levels of bias across datasets were assigned to the highest observed risk tier. This approach consolidates cross-dataset fairness assessments into a unified and interpretable framework, enabling targeted interventions to enhance equity in clinical predictions.

Table 17 summarizes the final set of *High Risk* features, which includes clinically significant predictors such as Age, BMI, LDL, and HbA1c Level—each of which either demonstrated substantial disparities or violated all nine fairness conditions across multiple datasets.

**Table 17.**
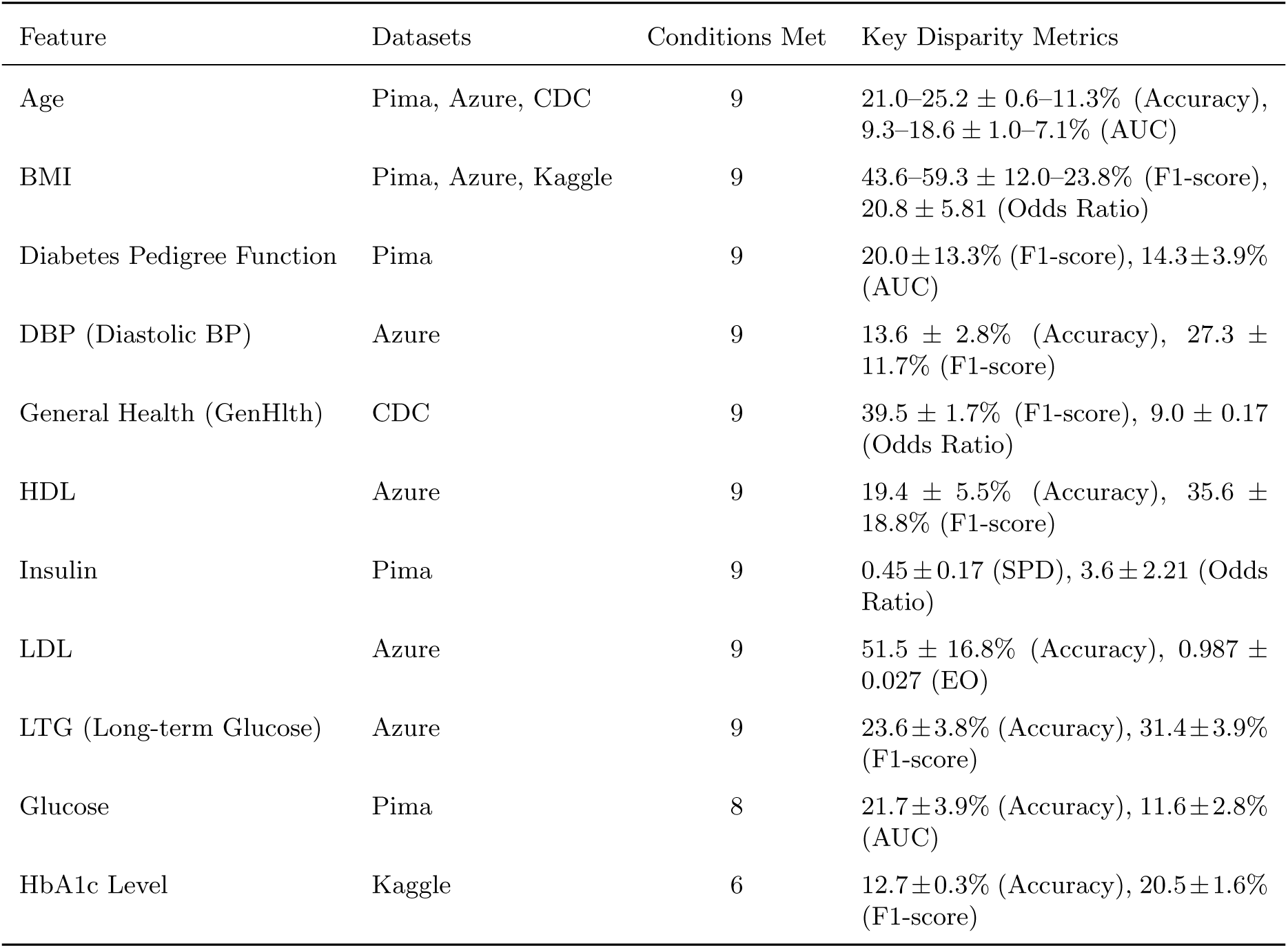
Summary of *High Risk* features identified across datasets based on revised fairness thresholds, sorted by Conditions Met in descending order.

The classification of *Blood Glucose Level* and *HbA1c Level* as *High Risk*, despite meeting only six fairness conditions in the Kaggle dataset, warrants further explanation. Although both fell short of the typical threshold of 8–9 conditions required for High Risk classification, their significant disparities—including a 16.1 *±* 0.6% accuracy disparity and 10.8 *±* 3.8% F1-score disparity for *Blood Glucose Level*, and a 12.7 *±* 0.3% accuracy disparity and 20.5 *±* 1.6% F1-score disparity for *HbA1c Level* —approached the severity observed in fully qualified High Risk features.

These disparities indicate that both features, as cornerstone clinical predictors of diabetes, can disproportionately affect prediction accuracy across demographic groups, potentially exacerbating inequities in high-stakes diagnostic settings. From a pathophysiological perspective, their complementary roles are critical: *Blood Glucose Level* reflects immediate glycemic status, whereas *HbA1c Level* captures long-term glucose control. These considerations collectively justified their elevation to High Risk, underscoring our commitment to prioritizing clinical relevance and patient safety in fairness evaluations.

It is also important to distinguish between the *Blood Glucose Level* feature in the Kaggle dataset and the *Glucose* feature in the Pima Indian Diabetes dataset. While *Blood Glucose Level* in Kaggle refers to a general, unspecified measurement of glucose concentration at a single time point—without information on testing conditions—*Glucose* in Pima specifically denotes plasma glucose concentration measured two hours after an oral glucose tolerance test (OGTT). This distinction is not merely semantic; the features differ in clinical meaning, data context, and diagnostic precision. As such, they must be treated as independent variables in both performance analysis and fairness assessment to avoid misleading generalizations across datasets.

Following the classification of *High Risk* features, we identified a set of *Medium Risk* features that violated between 6 and 8 fairness conditions and exhibited moderate disparities in model performance (e.g., accuracy between 10% and 20%, F1-score between 10% and 40%).

In the CDC dataset, the features *High Blood Pressure*, *Income*, and *Cholesterol Check* violated 8, 8, and 7 conditions respectively. While these features did not reach the disparity levels of High Risk predictors, they demonstrated non-trivial bias. For instance, *High Blood Pressure* showed a 14.5 *±* 0.3% accuracy disparity and an EOD of 0.137 *±* 0.009, whereas *Cholesterol Check* had an 11.2 *±* 0.6% accuracy disparity but did not exceed the High Risk threshold.

In the Kaggle dataset, *Blood Glucose Level* violated six fairness conditions and exhibited a 16.1 *±* 0.6% accuracy disparity and 10.8 *±* 3.8% F1-score disparity. Although noteworthy, these disparities fell short of the thresholds used for High Risk classification.

While impactful, these features were classified as Medium Risk due to their lower condition counts and moderate disparity levels when compared to High Risk features. Table 18 summarizes these *Medium Risk* features.

**Table 18.** Summary of *Medium Risk* features identified across datasets based on fairness violations.

| Feature | Datasets | Conditions Met | Key Disparity Metrics |
| --- | --- | --- | --- |
| High Blood Pressure | CDC | 8 | $14.5 \pm 0.3\%$ (Accuracy), $0.137 \pm 0.009$ (EOD) |
| Income | CDC | 8 | $12.2 \pm 1.3\%$ (Accuracy), $0.209 \pm 0.010$ (EOD) |
| Ment-health | CDC | 8 | $4.8 \pm 0.6\%$ (Accuracy), $10.6 \pm 1.3\%$ (F1-score), $0.150 \pm 0.015$ (EOD) |
| Phys-activity | CDC | 8 | $6.0 \pm 0.2\%$ (Accuracy), $8.4 \pm 0.7\%$ (F1-score), $0.109 \pm 0.012$ (EOD) |
| Cholesterol Check | CDC | 7 | $11.2 \pm 0.6\%$ (Accuracy), $14.6 \pm 3.3\%$ (F1-score) |
| Blood Glucose Level | Kaggle | 6 | $16.1 \pm 0.6\%$ (Accuracy), $10.8 \pm 3.8\%$ (F1-score) |

Finally, we identified a set of *Low Risk* features that violated fewer than six fairness conditions and exhibited only mild disparities in model performance (e.g., accuracy disparity ¡ 10%, F1-score disparity ¡ 10%).

In the Kaggle dataset, the features *Heart Disease*, *Hypertension*, and *Smoking History*, and in the CDC dataset, the feature *Smoking*, each violated only four or five conditions and demonstrated minimal disparities. For example, *Smoking* (CDC) showed an accuracy disparity of 3.5 *±* 0.1%, while *Smoking History* (Kaggle) had an accuracy disparity of 4.8 *±* 0.4%.

In the Pima dataset, the feature *Pregnancies*, which violated five fairness conditions, exhibited an accuracy disparity of 9.0 *±* 3.6%, placing it within the Low Risk tier as well.

We evaluated *Smoking History* (Kaggle) and *Smoking* (CDC) separately due to their distinct definitions: *Smoking History* is a multi-class categorical variable capturing various smoking statuses (e.g., former, current, never), while *Smoking* is a binary feature indicating current smoking status.

The low disparity levels and limited fairness violations support the classification of these features as Low Risk. Table 19 summarizes the identified Low Risk features.

**Table 19.** Summary of *Low Risk* features identified across datasets based on fairness violations.

| Feature | Datasets | Conditions Met | Key Disparity Metrics |
| --- | --- | --- | --- |
| Heart Disease | Kaggle | 5 | $8.0\% \pm 0.6\%$ (Accuracy), $0.18 \pm 0.017$ (SPD) |
| Hypertension | Kaggle | 5 | $6.7\% \pm 0.7\%$ (Accuracy), $0.15 \pm 0.006$ (SPD) |
| Pregnancies | Pima | 5 | $9.0\% \pm 3.6\%$ (Accuracy), $0.123 \pm 0.047$ (F1-score) |
| Smoking History | Kaggle | 5 | $4.8\% \pm 0.4\%$ (Accuracy), $0.072 \pm 0.021$ (F1-score) |
| Smoking | CDC | 3 | $3.5\% \pm 0.1\%$ (Accuracy), $0.72 \pm 0.01$ (DI) |

### Smart User Interface

This study evaluates the performance of the Smart User Interface (Smart UI) in enhancing fairness in diabetes prediction. Unlike prior analyses based solely on structured datasets (Kaggle Diabetes Prediction, Pima Indian Diabetes, Azure Open Diabetes, and CDC Diabetes Health Indicators), we simulate a real-world clinical environment using the Diabetes Dataset 2019 from Kaggle [62], which includes diverse and novel features.

This dataset comprises 994 instances with a wide range of demographic (e.g., age, gender), clinical (e.g., BMI, blood pressure, family history), and lifestyle-related (e.g., sleep, junk food consumption, physical activity) features. Its heterogeneity enables robust disparity analysis across age, gender, and socioeconomic subgroups, offering a practical scenario for testing the Smart UI’s adaptability to new predictors while leveraging interventions derived from previous datasets.

In our simulation, the Smart UI integrates data from multiple sources: wearable devices (e.g., sleep quality), user-provided responses (e.g., “How often do you eat junk food?”), and clinical notes extracted using natural language processing (NLP). Missing values are handled through iterative imputation, and a Random Forest classifier is used for prediction.

The interface presents fairness-aware outputs through subgroup-sensitive alerts, interactive dashboards, heatmaps, and plain-language summaries designed for both clinicians and patients. **Unlike conventional interfaces that passively present model outputs, the Smart UI enables real-time identification of potential subgroup biases and offers redesigned data entry flows to mitigate them.**

Our results show that the Smart UI effectively reduces disparities while maintaining predictive accuracy. For instance, the Equal Opportunity Difference (EOD) for *Age* decreased from 0.353 to 0.255 (Table 20), while overall accuracy remained stable at 92.3% *±* 1.2%. These findings position the Smart UI as a scalable, adaptable, and equitable decision-support tool for diabetes care.

**Table 20.** Fairness and performance metrics for the Age feature: Baseline vs. Smart UI on the Diabetes 2019 dataset. Improvements are marked with ^∗^.

| Metric | Baseline (Mean ± SD) | Smart UI (Mean ± SD) |
| --- | --- | --- |
| Regression Significance | $2.84 \times 10^{-19} \pm 0.00$ | $5.27 \times 10^{-18} \pm 3.49 \times 10^{-18} *$ |
| Accuracy Sensitivity (%) | $16.67 \pm 0.00$ | $13.25 \pm 3.84 *$ |
| AUC Sensitivity (%) | $13.26 \pm 0.29$ | $7.44 \pm 1.22 *$ |
| F1-Score Sensitivity (%) | $29.03 \pm 0.00$ | $22.24 \pm 5.19 *$ |
| Statistical Parity Diff. (SPD) | $0.189 \pm 0.009$ | $0.212 \pm 0.026$ |
| Disparate Impact (DI) | $0.371 \pm 0.011$ | $0.358 \pm 0.062$ |
| Equal Opportunity Diff. (EOD) | $0.353 \pm 0.000$ | $0.255 \pm 0.065 *$ |
| Equalized Odds (EO) | $0.353 \pm 0.000$ | $0.255 \pm 0.065 *$ |
| Odds Ratio (OR) | $2.70 \pm 0.08$ | $2.86 \pm 0.37$ |
| Overall Accuracy (%) | $90.52 \pm 0.28$ | $91.83 \pm 1.85 *$ |
| Overall F1-Score (%) | $80.35 \pm 0.47$ | $83.11 \pm 3.89 *$ |

### Rationale for Feature Categorization

To evaluate the adaptability of the Smart User Interface (Smart UI), we assessed its performance on the Diabetes Dataset 2019, which includes four novel features not present in prior datasets: stress, prediabetes (pdiabetes), urination frequency, and quality sleep (soundsleep). These features were categorized into appropriate fairness risk tiers by drawing analogies to similar features listed in the updated high, medium, and low risk tables.

Stress was classified as a *Medium Risk* feature due to its behavioral similarity to mental health, which exhibited moderate disparities in the CDC dataset—violating eight fairness conditions and showing a 10.6% *±* 1.3% F1-score disparity. Clinical literature further supports stress as a determinant of diabetes outcomes across population subgroups [4].

Prediabetes was assigned to the *High Risk* tier based on its physiological parallel to HbA1c, a feature already categorized as high risk due to its 20.5% *±* 1.6% F1-score disparity despite meeting only six fairness conditions [23].

In contrast, urination frequency and quality sleep were classified as *Low Risk* features given their symptomatic or lifestyle-related nature. They align with features such as smoking and physical activity, which showed minimal disparities. For instance, smoking in the CDC dataset violated only three conditions and had an accuracy disparity of 3.5% *±* 0.1%.

To validate consistency in model behavior, we assessed Smart UI’s performance on shared features such as age and BMI across both the Kaggle and Diabetes 2019 datasets. As summarized in Table 17, both features are considered high risk in all baseline datasets. Age showed an accuracy disparity ranging from 21% to 25%, and BMI demonstrated F1-score disparities between 43% and 59%. In our evaluation, the Smart UI reduced the accuracy disparity for age from 6.0% to 3.8%, with similar improvements observed for BMI (Tables 20 and 21).

**Table 21.**
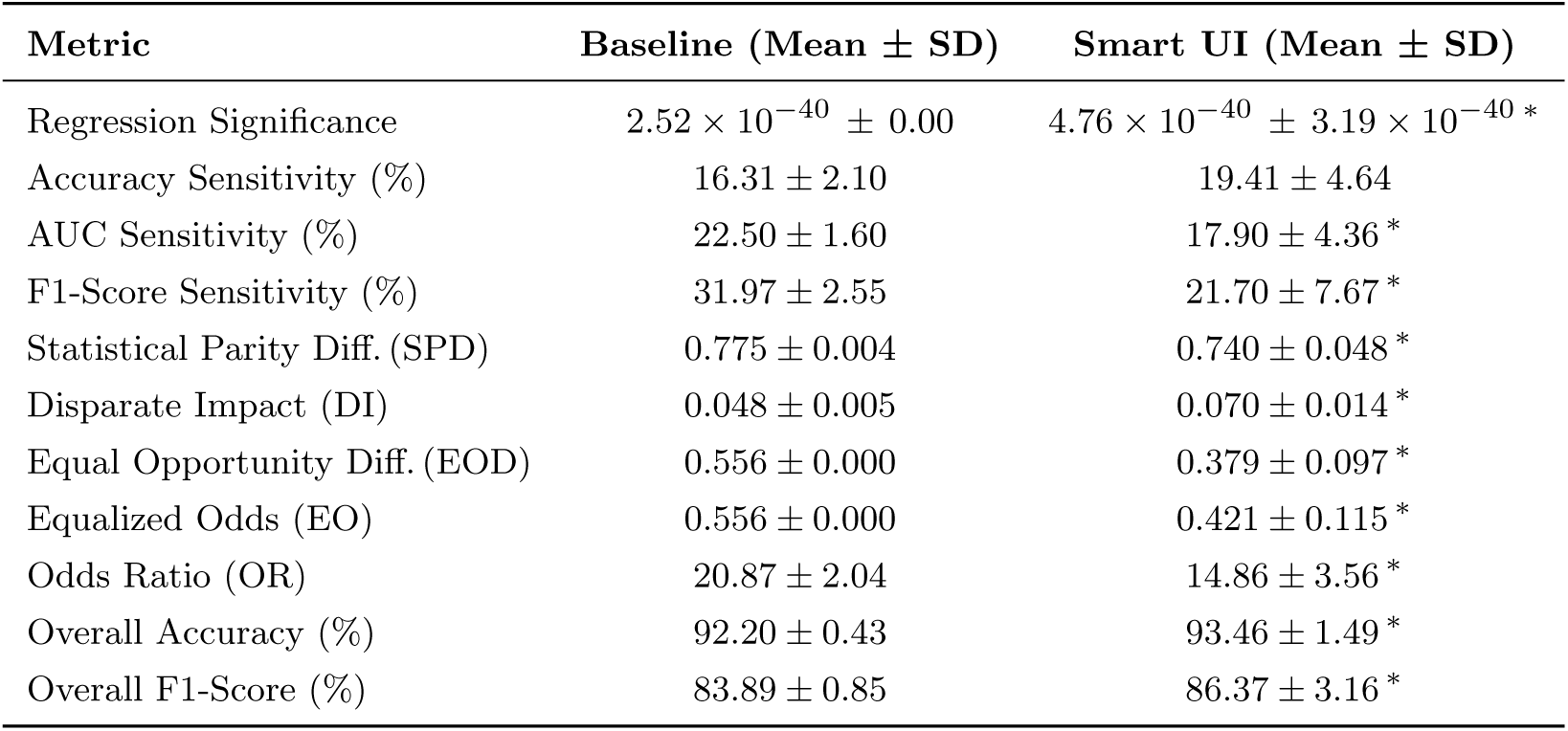
Fairness and performance metrics for the BMI feature: Baseline vs. Smart UI on the Diabetes 2019 dataset. Improvements are marked with ^∗^.

For a fair comparison with the baseline model, five commonly studied features—age, gender, BMI, blood pressure, and number of pregnancies—were selected from the Diabetes Dataset 2019. This selection was based on their clinical relevance, prevalence in standard datasets (e.g., Pima and CDC), and potential for fairness violations. According to the updated risk categorization, age and BMI remain high risk, while number of pregnancies is classified as low risk.

To simulate real-world data availability, only 70% of the contextual features (stress, prediabetes, urination frequency, and quality sleep) were retained, reflecting challenges such as inconsistent wearable usage or incomplete clinical records. This proportion was chosen empirically to represent realistic data completeness. Missing values were imputed using the Iterative Imputer method, and all features were normalized.

This experimental configuration specifically tested Smart UI’s ability to mitigate disparities in high-risk features such as age and BMI, even when contextual information was partially missing. For instance, quality sleep may help refine age-related predictions when subgroup bias is detected.

### Management of High-Risk Features

According to Table 17, age and BMI were identified as High Risk due to significant fairness disparities (e.g., 6.0% accuracy disparity for age). To evaluate their management, we compared fairness metrics between the baseline model (using raw data) and the Smart UI (using contextual adjustments). The UI leverages contextual features to refine predictions and reduce disparities.

The results presented in Tables 20 and 21 were obtained using ten-fold crossvalidation and a Random Forest model with 100 estimators. For the Age feature, Smart UI increased overall accuracy from 90.5% to 91.8% and F1-score from 80.4% to 83.1%, while equal opportunity difference (EOD) decreased from 0.353 to 0.255, demonstrating both enhanced predictive performance and a notable reduction in subgroup disparity.

### Categorization of Additional Features

The four additional features—stress, prediabetes (pdiabetes), urination frequency (urinationfreq), and quality sleep (soundsleep)—were categorized based on analogies to established features in our final risk framework.

Medium risk features are typically behavioral or psychological in nature, introducing disparities through social determinants such as gender or socioeconomic status. Accordingly, *stress* was classified as medium risk, due to its similarity to *mental health* in Table 18. Likewise, *prediabetes* was assigned to the medium risk tier, reflecting its metabolic role and conceptual similarity to *BMI*, a known high-risk feature.

Conversely, *urination frequency* and *quality sleep* were categorized as low risk due to their symptomatic or lifestyle-based nature. *Urination frequency*, akin to *smoking* in Table 19, is associated with minimal bias due to its subjective and reactive characteristics. *Quality sleep* was similarly placed in the low-risk group, reflecting the pattern of lifestyle features with protective or indirect effects, such as *heart disease* and *smoking history*.

### Visualization Strategy for Medium-Risk Features

The feature *Stress*, categorized as Medium Risk due to its behavioral similarity to mental health, is represented using a stratified line chart (Figure 9). This variable is ordinal with four levels: *not at all*, *sometimes*, *always*, and *very often*. The x-axis denotes the stress levels, and the y-axis indicates the mean predicted risk of diabetes. Distinct lines represent demographic subgroups, such as gender or socioeconomic status (SES). The visualization clearly illustrates that as stress level increases, predicted risk also rises. Moreover, the Smart UI version displays a visible reduction in equal opportunity difference (EOD) compared to the baseline model, highlighting its fairness-enhancing potential.

**Fig. 9.**
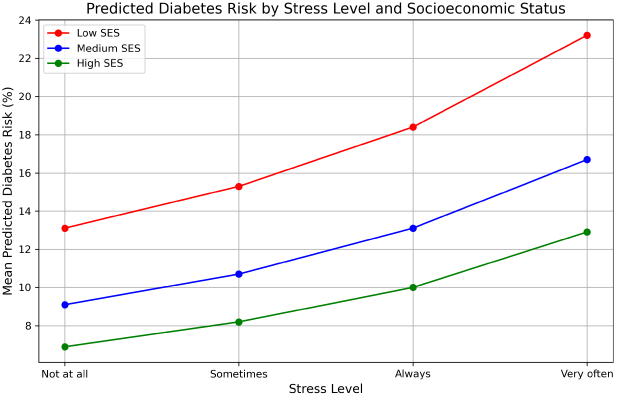
Stratified line chart showing predicted diabetes risk across stress levels, segmented by SES groups.

The binary feature *Prediabetes (pdiabetes)*, also categorized as Medium Risk due to its metabolic association with features like BMI, is visualized using a grouped bar chart (Figure 10). This chart compares the mean predicted diabetes risk between individuals with and without prediabetes. Bars are stratified by sensitive attributes such as gender or access to healthcare. The resulting pattern reveals a reduction in statistical parity difference (SPD) when contextual adjustments provided by the Smart UI are applied.

**Fig. 10.**
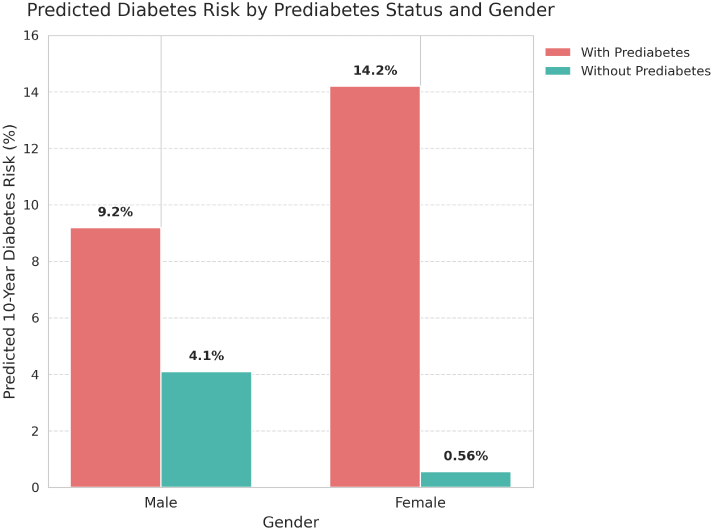
Grouped bar chart comparing predicted diabetes risk for individuals with and without prediabetes, stratified by gender.

Both charts are dynamically generated within the Smart User Interface in response to the entry of relevant data, such as new stress levels or prediabetes status. This realtime feedback allows clinicians and researchers to identify emerging disparities, verify input accuracy, or request additional clinical information where needed. By enabling continuous monitoring and interpretation of medium-risk features, the Smart UI not only maintains predictive accuracy but also actively mitigates bias, supporting fairer clinical decision-making processes.

### Monitoring Strategy for Low-Risk Features

The feature *Urinationfreq*, classified as Low Risk due to its limited symptomatic impact on diabetes prediction, is a binary variable with levels *low* and *high*. This feature is monitored via a real-time alert in the Smart User Interface, which compares the mean predicted diabetes risk between the two groups. The alert message clearly states that patients with high urination frequency show a higher predicted risk compared to those with low frequency. If the observed gap exceeds a predefined fairness threshold, a red warning card labeled “ **ALERT: Urination Frequency**” is triggered. This alert explicitly reports the gap size, indicates the threshold exceedance, and suggests specific clinical actions such as reviewing patient input data or requesting a glucose test.

Similarly, the feature *Soundsleep*, also categorized as Low Risk due to its protective and lifestyle-related nature, is a continuous variable ranging from 0 to 10. It is monitored through a dynamic alert that tracks the maximum risk gap across binned sleep categories (e.g., 0–3, 4–6, 7–10). If the gap remains within acceptable range, a non-intrusive notification card labeled “ **NOTICE: Sleep Quality**” is shown, indicating that the system is still monitoring and no immediate action is required.

These alert cards are displayed in real time for clinicians, researchers, and patients within the Smart UI whenever new values for *Urinationfreq* or *Soundsleep* are entered, allowing the system to instantly flag fairness-relevant disparities. A visual example of these real-time alerts is illustrated in Figure 11, highlighting how the Smart UI communicates risks and suggests corrective actions through interpretable, actionable messages.

**Fig. 11.**
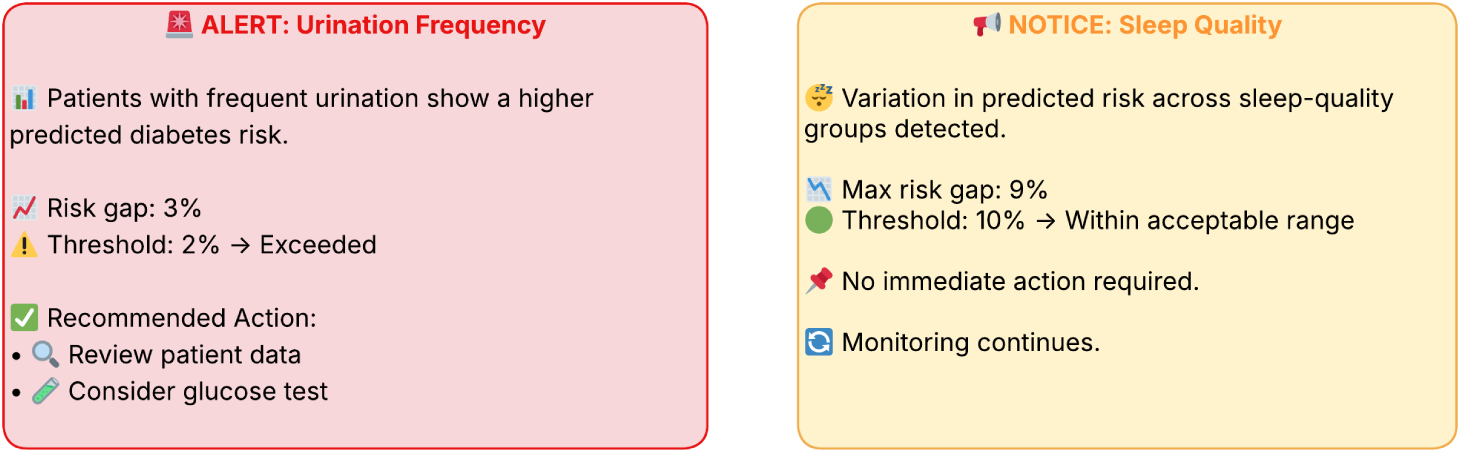
Smart UI alert cards for Low Risk features. Left: Alert for *Urinationfreq* triggered when risk disparity exceeds the fairness threshold. Right: Notice for *Soundsleep* indicating monitoring status when disparity remains within acceptable range.

In a clinical scenario, if the system detects that the predicted diabetes risk for patients with high sleep quality and limited healthcare access is unexpectedly higher than for a comparable group with better access, an alert is activated. This encourages the clinician to verify data integrity or consider requesting a complementary test such as a blood glucose assessment. In this way, the Smart UI enables the detection and mitigation of subtle biases even in Low Risk features, supporting fairer and more informed decisions for vulnerable populations.

### Limitations and Future Directions

This study was conducted in a simulated environment and assumes high-quality data collection and seamless interaction with the Smart User Interface (Smart UI). The natural language processing (NLP) extraction module was modeled with a fixed success probability of 70%; however, real-world performance of clinical NLP systems may vary considerably depending on the quality, structure, and complexity of the text. Previous research indicates that the accuracy of state-of-the-art clinical NLP tools ranges from 60% to 90% depending on the task and dataset [63]. In addition, the dynamic nature of clinical data input may encounter challenges such as incomplete electronic health records (EHRs), entry errors, or limited access to wearable devices, all of which can introduce data noise or sparsity and reduce predictive robustness.

Another limitation of the study is the use of a single dataset for Smart UI validation. Although the Diabetes Dataset 2019 provides a rich variety of features, it does not represent the full heterogeneity of global populations or clinical settings. Furthermore, no usability testing has yet been conducted with real users (e.g., clinicians or patients), and the current interface design is based on theoretical assumptions regarding user behavior and needs rather than empirical feedback.

Future work will focus on real-world deployment of the Smart UI in clinical environments to collect live, user-generated data. Validation across geographically and socioeconomically diverse populations will be prioritized to evaluate generalizability and fairness. In addition, structured usability studies will be conducted with healthcare professionals and patients to assess and optimize user interaction, interpretability, and actionability of the interface. These efforts will be complemented by the exploration of adaptive learning mechanisms—such as reinforcement learning and feedback-driven interface refinement—to improve resilience against incomplete or noisy data and enhance long-term system performance.

These directions are aligned with the manuscript’s broader vision for scalable, user-centered, and fairness-aware AI in healthcare, and advance the goal of safe, equitable, and actionable diabetes prediction in real-world clinical practice.

## Discussion

This study introduces a novel Smart User Interface (Smart UI) framework that redefines fairness intervention in AI-driven diabetes prediction systems. In contrast to prevailing strategies that emphasize bias mitigation at the data or algorithmic level, our approach shifts the focus to the point of interaction—where humans and machines meet. By embedding fairness-aware logic directly into the interface, we enable actionable, context-sensitive decision support that empowers clinicians without requiring modifications to the underlying models or data pipelines.

The Smart UI stands apart from existing tools such as Fairlearn and Aequitas, which provide retrospective, developer-focused fairness audits. These tools, while essential for initial evaluations, are often detached from clinical workflows and offer limited utility at the moment of care. Our interface, by comparison, is designed for real-time deployment within electronic health record (EHR) systems and supports end-users—clinicians, patients, and researchers—with intuitive, role-specific views. It dynamically visualizes equity risks, generates contextual alerts, and surfaces tailored summaries, thereby converting abstract bias metrics into interpretable clinical insights. Evaluation across four demographically diverse datasets—Kaggle, Pima Indian, Azure, and CDC Diabetes—demonstrated the framework’s effectiveness in reducing subgroup disparities. For example, the equal opportunity difference (EOD) for age was reduced from 0.35 to 0.25, and for BMI from 0.56 to 0.38, indicating meaningful improvements in equity without compromising model performance. These results underscore the potential of interface-level interventions as a model-agnostic and low-friction strategy for promoting fairness in clinical AI applications.

A key innovation of our approach lies in its multidimensional integration of patient context. The Smart UI consolidates structured EHR data with real-time signals from wearable devices and unstructured clinical notes via natural language processing. This generates a dynamic patient profile that enables more informed, personalized, and equitable decision-making. Such design is especially pertinent to diabetes care, where socio-behavioral factors—often overlooked by traditional models—play a critical role in outcomes. By presenting this complexity in a clinically usable format, the interface bridges the gap between fairness research and real-world healthcare delivery.

The open-source, modular architecture of the Smart UI further enhances its adaptability and collaborative potential. Developers can extend components, institutions can tailor workflows, and research communities can build on shared foundations. This positions the framework not just as a technical contribution, but as a platform for collective innovation in equitable AI.

Nevertheless, several limitations must be addressed in future work. First, our evaluations were conducted using publicly available and simulated datasets, which may not fully capture the heterogeneity of global clinical populations. To strengthen external validity, future studies should involve underrepresented regions—such as sub-Saharan Africa and Southeast Asia—and conduct multi-center clinical trials to assess usability, impact, and generalizability. Second, although designed with usability in mind, integrating Smart UI into clinical workflows may still pose practical challenges. We are actively collaborating with healthcare providers for iterative co-design and real-time usability testing. Third, patient trust is a cornerstone of ethical AI. While patients do not directly interact with the UI, we address privacy and inclusivity through robust encryption and culturally sensitive content. Future work will expand on these efforts by incorporating patient advisory boards into the design of communication outputs.

In summary, this study reframes fairness not merely as a mathematical objective but as a human-centered imperative. By addressing equity at the interface level, we offer a practical, replicable, and ethical pathway for improving diagnostic fairness. The Smart UI is not simply a tool—it is a paradigm shift in how AI fairness can be realized in clinical contexts. Future directions include real-world implementation, broader cross-cultural validation, and the development of adaptive features that align with evolving clinical standards and regulatory frameworks. Through this lens, we move closer to a healthcare system that is not only intelligent, but just.

## Author Contributions

All authors made substantial contributions to the concept, design, and execution of this study.

## Data Availability

No new data are associated with this study.

https://www.cdc.gov/brfss/index.html

https://github.com/Azure/OpenDatasets

https://www.kaggle.com/datasets/uciml/pima-indians-diabetes-database

https://www.kaggle.com/datasets/iammustafatz/diabetes-prediction-dataset

## References

[1] International Diabetes Federation: Idf diabetes atlas. Diabetes Research and Clinical Practice 183, 109119 (2021) 10.1016/j.diabres.2021.109119

[2] LaVeist, T.A., Thorpe, R.J., Galarraga, J.E., Bower, K.M., Gary-Webb, T.L.: Environmental and socio-economic factors as contributors to racial disparities in diabetes prevalence. American Journal of Public Health 104(S4), 659–666 (2014) 10.2105/AJPH.2013.301774

[3] Zamora-Kapoor, A., Sinclair, K., Braun, K., Lambert, W.: Epidemiology of type 2 diabetes in indigenous communities in the united states. Current Diabetes Reports 21(9), 1–8 (2021) 10.1007/s11892-021-01426-6

[4] Hill-Briggs, F., Adler, N.E., Berkowitz, S.A., Chin, M.H., Gary-Webb, T.L., Navas-Acien, A., Thornton, P.L., Haire-Joshu, D.: Social determinants of health and diabetes: A scientific review. Diabetes Care 44(1), 258–279 (2021) 10.2337/dci20-0053

[5] Mohsen, F., Al-Saadi, B., Abdi, N., Khan, S., Shah, Z.: Artificial intelligence-based methods for predicting the risk of type 2 diabetes mellitus: A systematic review. npj Digital Medicine 6(1), 1–16 (2023) 10.1038/s41746-023-00933-5

[6] Obermeyer, Z., Powers, B., Vogeli, C., Mullainathan, S.: Dissecting racial bias in an algorithm used to manage the health of populations. Science 366(6464), 447–453 (2019) 10.1126/science.aax2342

[7] Vokinger, K.N., Feuerriegel, S., Kesselheim, A.S.: Mitigating bias in machine learning for medicine. npj Digital Medicine 6(1), 1–8 (2023) 10.1038/s41746-023-00858-z

[8] iammustafatz: Diabetes Prediction Dataset. Kaggle. Accessed: 22 April 2025 (2023)

[9] mathchi: Diabetes Data Set. Kaggle. Accessed: 22 April 2025 (2019)

[10] Microsoft: Azure Open Datasets: Diabetes. Microsoft. Accessed: 22 April 2025 (2020)

[11] Bardenheier, B. H. and Bullard, K. M. and Caspersen, C. J. and Cheng, Y. J. and Gregg, E. W. and Geiss, L. S.: CDC Diabetes Health Indicators Dataset. Mendeley Data. Accessed: 22 April 2025 (2020)

[12] Lee, S.-H., Park, S.Y., Kim, Y.J., Park, J.S.: Lifestyle factors and the risk of type 2 diabetes: A review. Diabetologia 62(10), 1777–1787 (2019) 10.1007/s00125-019-4955-9

[13] Mozaffarian, D., Kamineni, A., Carnethon, M., Djoussé, L., Mukamal, K.J., Siscovick, D.: Lifestyle risk factors and new-onset diabetes mellitus in older adults. Archives of Internal Medicine 170(8), 698–705 (2010) 10.1001/archinternmed.2010.21

[14] American Diabetes Association: Standards of medical care in diabetes— 2024. Diabetes Care 47(Supplement 1), 1–321 (2024) 10.2337/dc24-SINT

[15] National Institute for Health and Care Excellence: Type 2 diabetes in adults: Management (NG28). https://www.nice.org.uk/guidance/ng28. Accessed: 22 April 2025 (2015)

[16] Mehrabi, N., Morstatter, F., Saxena, N., Lerman, K., Galstyan, A.: A Survey on Bias and Fairness in Machine Learning. arXiv preprint arXiv:1908.09635. Accessed: 22 April 2025 (2019). https://arxiv.org/abs/1908.09635

[17] Chouldechova, A.: Fair prediction with disparate impact: A study of bias in recidivism prediction instruments. Big Data 5(2), 153–163 (2017) 10.1089/big.2016.0047

[18] Grgic-Hlaca, N., Zafar, M.B., Gummadi, K.P., Weller, A.: The case for process fairness in learning: Feature selection for fair decision making. In: NIPS Symposium on Machine Learning and the Law, pp. 1–4 (2016)

[19] Fiaz, M., Javed, A., Saleem, M.A., Ijaz, H., Khan, S.: Ethical frameworks for machine learning in sensitive healthcare applications. Frontiers in Medicine 10, 10789841 (2023) 10.3389/fmed.2023.10789841

[20] Lekadir, K., Frangi, A.F., Porras, A.R., Glocker, B., Cintas, C., Langlotz, C.P., Weicken, E., Asselbergs, F.W., Prior, F., Collins, G.S., Kaissis, G., Tsakou, G., Buvat, I., Kalpathy-Cramer, J., Mongan, J., Schnabel, J.A., Kushibar, K., Riklund, K., Marias, K., Amugongo, L.M., Fromont, L.A., Maier-Hein, L., Cerdá-Alberich, L., Martí-Bonmatí, L., Cardoso, M.J., Bobowicz, M., Shabani, M., Tsiknakis, M., Zuluaga, M.A., Fritzsche, M.-C., Camacho, M., Linguraru, M.G., Wenzel, M., De Bruijne, M., Tolsgaard, M.G., Goisauf, M., Cano badía, M., Papanikolaou, N., Lazrak, N., Pujol, O., Osuala, R., Napel, S., Colantonio, S., Joshi, S., Klein, S., Aussó, S., Rogers, W.A., Salahuddin, Z., Starmans, M.P.A.: Future-ai: International consensus guideline for trustworthy and deployable artificial intelligence in healthcare. BMJ 388, 081554 (2025) 10.1136/bmj-2024-081554

[21] Gianfrancesco, M.A., Tamang, S., Yazdany, J., Schmajuk, G.: Potential biases in machine learning algorithms using electronic health record data. JAMA Internal Medicine 179(11), 1544–1547 (2019) 10.1001/jamainternmed.2018.3763

[22] Li, Y., Schoufour, J., Wang, D.D., Dhana, K., Pan, A., Liu, X., Song, M., Liu, G., Shin, H.J., Sun, Q., Al-Shaar, L., Wang, M., Rimm, E.B., Hertzmark, E., Stampfer, M.J., Willett, W.C., Franco, O.H., Hu, F.B.: Combined lifestyle factors and risk of incident type 2 diabetes and prognosis among individuals with type 2 diabetes: A systematic review and summary of meta-analysis studies. Diabetologia 63(1), 1–15 (2019) 10.1007/s00125-019-04985-9

[23] Chung, H., Ko, H., Kang, W.S., Yoon, K., Lee, H.: Machine learning models for data-driven prediction of diabetes by lifestyle type. International Journal of Environmental Research and Public Health 19(22), 15027 (2022) 10.3390/ijerph192215027

[24] Patel, V.A., Goldstein, B.A., Shah, S.H.: Predicting changes in glycemic control among adults with prediabetes from activity patterns collected by wearable devices. npj Digital Medicine 4, 541 (2021) 10.1038/s41746-021-00541-1

[25] Lee, S., et al.: Machine learning models for data-driven prediction of diabetes by lifestyle type. International Journal of Environmental Research and Public Health 19(22), 15027 (2022) 10.3390/ijerph192215027

[26] Borrell, L.N., Dallo, F.J., White, K.: Education and diabetes in a racially and ethnically diverse population. American Journal of Public Health 96(9), 1637– 1642 (2006) 10.2105/AJPH.2005.072884

[27] Solanki, J.D., Makwana, A.H., Mehta, H.B., Gokhale, P.A., Shah, C.J.: Body composition in type 2 diabetes: Change in quality and not just quantity that matters. International Journal of Preventive Medicine 6, 122 (2015) 10.4103/2008-7802.172376

[28] Yoshimura, E., et al.: Effects of meal timing on postprandial glucose metabolism and blood metabolites in healthy adults. Nutrients 10(11), 1763 (2018) 10.3390/nu10111763

[29] Adkison, J.D., Chung, P.: Implementing continuous glucose monitoring in clinical practice. Family Practice Management 28(2), 7–14 (2021)

[30] Crofts, C., et al.: Postprandial insulin assay as the earliest biomarker for diagnosing pre-diabetes, type 2 diabetes and increased cardiovascular risk. Open Heart 4(2), 000656 (2017) 10.1136/openhrt-2017-000656

[31] Barter, P.J., et al.: Effect of statins on hdl-c: a complex process unrelated to changes in ldl-c: analysis of the voyager database. Journal of Lipid Research 51(6), 1546–1554 (2010) 10.1194/jlr.M002709

[32] Lindström, J., Tuomilehto, J.: The diabetes risk score: a practical tool to predict type 2 diabetes risk. Diabetes Care 26(3), 725–731 (2003) 10.2337/diacare.26.3.725

[33] Spruill, T.M.: Chronic psychosocial stress and hypertension. Current Hypertension Reports 12(1), 10–16 (2010) 10.1007/s11906-009-0084-8

[34] Chobanian, A.V., et al.: Seventh report of the joint national committee on prevention, detection, evaluation, and treatment of high blood pressure. Hypertension 42(6), 1206–1252 (2003) 10.1161/01.HYP.0000107251.49515.c2

[35] Kessler, R.C., et al.: The effects of chronic medical conditions on mental health. Archives of General Psychiatry 60(12), 1215–1223 (2003) 10.1001/archpsyc.60.12.1215

[36] Saleiro, P., et al.: Aequitas: A bias and fairness audit toolkit. arXiv (2018)

[37] Wexler, J., et al.: The what-if tool: Interactive probing of machine learning models. IEEE Transactions on Visualization and Computer Graphics 26(1), 56–65 (2020) 10.1109/TVCG.2019.2934619

[38] Weerts, H., et al.: Fairlearn: Assessing and improving fairness of ai systems. Journal of Machine Learning Research 24(1), 1–8 (2023)

[39] Wu, Z., et al.: A case study of integrating fairness visualization tools in machine learning education. CHI EA ’22, 356–17 (2022) 10.1145/3491101.3503568

[40] Saydah, S., et al.: Social determinants of health and diabetes: A scientific review. Diabetes Care 44(1), 258–279 (2021) 10.2337/dci20-0053

[41] Breck, E., et al.: Data Validation for Machine Learning. In: Proceedings of the 2nd SysML Conference (2019)

[42] Singh, A., et al.: Fairness drift detection for categorical features. Journal of Machine Learning Research 22(1), 1–30 (2021)

[43] Cabrera, A., et al.: Fairvis: Visual analytics for discovering intersectional bias in machine learning. Proceedings of the 2019 IEEE Conference on Visual Analytics Science and Technology (VAST) (2019) 10.1145/3290605.3300836

[44] Dodge, J., et al.: Explaining models: An empirical study of how explanations impact fairness judgment. Proceedings of the 2020 CHI Conference on Human Factors in Computing Systems (2020) 10.1145/3351095.3372858

[45] Ghosh, A., et al.: Faircanary: Rapid continuous explainable fairness. Proceedings of the 2022 AAAI/ACM Conference on AI, Ethics, and Society (2022)

[46] Lundberg, S.M., et al.: Explainable ai for trees: From local explanations to global understanding. Nature Machine Intelligence 1(5), 252–263 (2019) 10.1038/s42256-019-0138-9

[47] Amershi, S., et al.: Guidelines for human-ai interaction. Proceedings of the 2019 CHI Conference on Human Factors in Computing Systems (2019) 10.1145/3290605.3300233

[48] Kreimeyer, K., et al.: Natural language processing systems for clinical information. Journal of Biomedical Informatics (2017) 10.1016/j.jbi.2017.07.012

[49] Mickelson, R.S., et al.: Usability testing of a mobile health application. International Journal of Environmental Research and Public Health (2016) 10.3390/ijerph13090902

[50] Ratwani, R.M., et al.: Electronic health record usability. Journal of the American Medical Informatics Association (2015) 10.1093/jamia/ocv099

[51] Kim, S., Kim, J., Faiola, A.: Acceptability of a health care app with 3 user interfaces for older adults and their caregivers: Design and evaluation study. JMIR Aging 6, 10034616 (2023) 10.2196/41432

[52] Alharbi, A., Alosaimi, F., et al.: A model to improve user acceptance of e-services in healthcare systems based on technology acceptance model: an empirical study. Journal of Ambient Intelligence and Humanized Computing 14, 1–14 (2023) 10.1007/s12652-023-04601-0

[53] Johnson, C.M., et al.: A user-centered framework for redesigning health care interfaces. Journal of Biomedical Informatics (2015) 10.1016/j.jbi.2014.11.005

[54] Floridi, L., et al.: Ai4people—an ethical framework for a good ai society. Minds and Machines 28(4), 689–707 (2018) 10.1007/s11023-018-9482-5

[55] National Institute of Standards and Technology: Security and privacy controls for federal information systems and organizations (nist sp 800-53). Technical report, U.S. Department of Commerce (2013)

[56] Khalil, M., et al.: Security and privacy in mhealth applications. Journal of Medical Systems (2020) 10.1007/s10916-020-01564-3

[57] Koru UX: 50 Healthcare UX/UI Design Trends With Examples. Accessed: 2025-05-07. https://www.koruux.com/50-examples-of-healthcare-UI/

[58] Holzinger, A., et al.: What do we need to build explainable ai systems for health-care? Journal of Medical Internet Research (2017) 10.2196/jmir.6469

[59] Satyanarayanan, M., et al.: Edge analytics in the internet of things. IEEE Pervasive Computing 14(2), 24–31 (2015) 10.1109/MPRV.2015.32

[60] Berdahl, C.T., Mann, D.M., et al.: Improving provider adoption with adaptive clinical decision support surveillance: An observational study. JMIR Human Factors 6(1), 10245 (2019) 10.2196/10245

[61] Alharbi, A., Smith, J., et al.: What characteristics of clinical decision support system implementations lead to adoption for regular use? a scoping review. BMJ Health Care Informatics 31(1), 101046 (2024) 10.1136/bmjhci-2023-101046

[62] tigganeha4: Diabetes Dataset 2019. Kaggle. Accessed: 22 April 2025 (2020)

[63] Wu, H., Toti, G., Morley, K., Brock, A., Joshi, R., Devarakonda, M.: Clinicalbert: Modeling clinical notes and predicting hospital readmission. arXiv preprint arXiv:1904.05342 (2020)

